# Similarity and diversity of genetic architecture for complex traits between East Asian and European populations

**DOI:** 10.1101/2023.05.26.23290578

**Authors:** Jinhui Zhang, Shuo Zhang, Jiahao Qiao, Ting Wang, Ping Zeng

## Abstract

**Background:** Genome-wide association studies have detected a large number of single-nucleotide polymorphisms (SNPs) associated with complex traits in diverse ancestral groups. However, the trans-ethnic similarity and diversity of genetic architecture is not well understood currently.

**Results:** By leveraging summary statistics of 37 traits from East Asian (*N*_max_=254,373) or European (*N*_max_=693,529) populations, we first evaluated the trans-ethnic genetic correlation (*ρ_g_*) and found substantial evidence of shared genetic overlap underlying these traits between the two populations, with 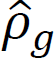 ranging from 0.53 (se=0.11) for adult-onset asthma to 0.98 (se=0.17) for hemoglobin A1c. However, 88.9% of the genetic correlation estimates were significantly less than one, indicating potential heterogeneity in genetic effect across populations. We next identified common associated SNPs using the conjunction conditional false discovery rate method and observed 21.7% of trait-associated SNPs can be identified simultaneously in both populations. Among these shared associated SNPs, 20.8% showed heterogeneous influence on traits between the two ancestral populations. Moreover, we demonstrated that population-common associated SNPs often exhibited more consistent linkage disequilibrium and allele frequency pattern across ancestral groups compared to population-specific or null ones. We also revealed population-specific associated SNPs were much likely to undergo natural selection compared to population-common associated SNPs.

**Conclusions:** Our study provides an in-depth understanding of similarity and diversity regarding genetic architecture for complex traits across diverse populations, and can assist in trans-ethnic association analysis, genetic risk prediction, and causal variant fine mapping.

## Background

Over the past few years, large-scale genome-wide association studies (GWASs) have convincingly detected a large number of genetic loci associated with a series of complex traits by casting single-nucleotide polymorphisms (SNPs) across the entire genome [1], generating novel biological knowledge and renovating diagnostic and treatment tools for diseases [2, 3]. However, existing GWASs have been heavily biased towards European (EUR) individuals, with 66.4% (=7,377/11,113, until 2023-4-25) of studies conducted in EUR but only a few in other populations [4]. As medical genomics studies have become increasingly large and diverse, acquiring insights into similarity and diversity of trait-associated SNPs among distinct populations and consequently the transferability of disease genetic risk is imperative in clinical translation [5].

It has been revealed that many trait-associated SNPs identified in GWASs of EUR ancestry can be replicated in other ethnic groups [2, 6–10], in the sense that they show significant association with consistent direction of genetic effects in non-EUR individuals, indicating complex traits enjoy common genetic components across diverse continental populations. For example, the trans-ethnic genetic correlation is 0.79 (se=0.04) for ulcerative colitis and 0.76 (se=0.04) for Crohn’s disease [11], 0.46 (se=0.06) for rheumatoid arthritis (RA) [12], 0.33 (se=0.03) for major depressive disorder [13], and 0.39 (se=0.15) for attention-deficit hyperactivity disorder between the EUR and East Asian (EAS) populations. However, these estimated trans-ethnic genetic correlation are in general significantly less than one in spite of being larger than zero [12, 14], which meanwhile suggests ancestral diversity.

Indeed, ancestry-relevant heterogeneity regarding varying allele frequency and linkage disequilibrium (LD) patterns is observed for some causal variants such that a significant association detected in one population is not readily identified in others [5, 15, 16]. Notable examples include a nonsense variant in *TBC1D4*, which confers muscle insulin resistance and increases risk for type 2 diabetes (T2D) and is common in Greenland but rare or absent in other populations [17], several common EAS-specific coding variants that influence blood lipids by exerting a protective effect against alcoholism [18, 19], and two loci associated with major depression that are more common in the Chinese population than EUR (i.e., 45% vs. 2% for rs12415800, and 28% vs. 6% for rs35936514) [20, 21]. As another example, multiple associated loci located within *PCSK9*, *APOA*, *APOC*, and *ABCA1* all play key roles in lipid genetics in both the EUR and African (AFR) Americans, yet the precise alleles within each locus differ substantially between the two populations, supporting the perspective that lipid-associated SNPs are largely shared across ancestral groups, but the allelic structure within a locus may be shaped by population history and thus exhibits considerable heterogeneity [22, 23].

Prior work has investigated the replicability of GWAS discoveries for some particular traits, displaying the similarity and diversity of associated SNPs across ancestral groups [7, 12, 24, 25]. However, those studies primarily focused on limited traits or a small set of significant loci [7, 24]; it is unknown whether their conclusions can be generalized to other traits or to genome-wide SNPs given the polygenic nature of many complex phenotypes. In addition, some of previous studies focused mainly on the trans-ethnic genetic correlation [12, 25, 26], which however only quantifies an overall genetic similarity across the whole genome and cannot characterize detailed association pattern for individual SNPs. A comprehensive genome-wide assessment of trans-ethnic similarity and diversity of genetic components for traits is challenging because our knowledge of genetic architecture within each population is not fully understood.

To fill in this critical knowledge gap mentioned above, here we obtained GWAS summary statistics of 37 complex traits from the EAS and EUR populations to perform a complete comparison of genetic similarity and diversity across the two populations. We first evaluated the trans-ethnic genetic correlation to quantify the extent of common genetic basis to which these traits shared [12]. Then, we identified population-specific and population-common trait-associated SNPs [27, 28]. Afterwards, we conducted the marginal genetic correlation analysis and heterogeneity test for these associated SNPs [25]. Finally, we evaluated how the LD and minor allele frequency (MAF) patterns varied among various types of SNPs and studied whether the genetic differentiation between ancestry groups can be explained via natural selection [7, 24]. We also assessed the genetic influence of associated SNPs on traits across populations by calculating a genetic risk score (GRS) [24, 29].

## Results

### Overview of the used statistical methods

We here give a brief overview of some important statistical methods employed in our analyses and showed more methodological descriptions in the Materials and Methods Section. Technical details of all used methods could be found in respective original papers. We here analyzed a total of 37 complex traits (10 binary and 27 continuous) using summary statistics obtained from EAS-only or EUR-only GWASs (**Table 1**).

**Table 1.** Summary information of complex traits analyzed in the present study

| trait | $\hat{h}_1^2$ ($se_1$) | $\hat{h}_2^2$ ($se_2$) | $P_{\Delta h^2}$ | $\hat{\rho}_g$ ($se$, $P_\rho$) | trait | $\hat{h}_1^2$ ($se_1$) | $\hat{h}_2^2$ ($se_2$) | $P_{\Delta h^2}$ | $\hat{\rho}_g$ ($se$, $P_\rho$) |
| --- | --- | --- | --- | --- | --- | --- | --- | --- | --- |
| SCZ | 0.360 (0.020) | 0.364 (0.015) | $8.73 \times 10^{-1}$ | **0.74 (0.10, $1.33 \times 10^{-2}$)** | TG | 0.117 (0.037) | 0.209 (0.045) | $1.14 \times 10^{-1}$ | - |
| RA | 0.143 (0.034) | 0.115 (0.016) | $4.56 \times 10^{-1}$ | **0.70 (0.14, $2.65 \times 10^{-2}$)** | HbA1c | 0.073 (0.014) | 0.039 (0.006) | **$2.56 \times 10^{-2}$** | 0.98 (0.17, $9.25 \times 10^{-1}$) |
| AF | 0.095 (0.027) | 0.020 (0.003) | **$5.77 \times 10^{-3}$** | - | eGFR | 0.071 (0.007) | 0.058 (0.004) | $1.07 \times 10^{-1}$ | **0.83 (0.05, $4.42 \times 10^{-4}$)** |
| T2D | 0.066 (0.004) | 0.041 (0.002) | **$2.27 \times 10^{-8}$** | 0.93 (0.04, $5.89 \times 10^{-2}$) | AAM | 0.081 (0.008) | 0.125 (0.006) | **$1.08 \times 10^{-5}$** | - |
| COA | 0.015 (0.003) | 0.043 (0.006) | **$3.00 \times 10^{-5}$** | **0.57 (0.09, $1.66 \times 10^{-6}$)** | ANM | 0.082 (0.013) | 0.132 (0.013) | **$6.54 \times 10^{-3}$** | **0.66 (0.09, $2.34 \times 10^{-4}$)** |
| AOA | 0.015 (0.003) | 0.030 (0.003) | **$4.07 \times 10^{-4}$** | **0.53 (0.11, $1.21 \times 10^{-5}$)** | PLT | 0.011 (0.014) | 0.201 (0.018) | **$7.95 \times 10^{-17}$** | **0.84 (0.07, $2.51 \times 10^{-2}$)** |
| AD | 0.008 (0.002) | 0.040 (0.015) | **$3.45 \times 10^{-2}$** | - | RBC | 0.085 (0.010) | 0.151 (0.017) | **$8.19 \times 10^{-4}$** | 0.92 (0.06, $1.39 \times 10^{-1}$) |
| BRC | 0.053 (0.030) | 0.106 (0.010) | $9.38 \times 10^{-2}$ | - | MCV | 0.132 (0.018) | 0.227 (0.032) | **$9.67 \times 10^{-3}$** | 0.87 (0.07, $7.48 \times 10^{-2}$) |
| IS | 0.010 (0.002) | 0.008 (0.001) | $3.71 \times 10^{-1}$ | 0.89 (0.20, $5.69 \times 10^{-1}$) | HCT | 0.052 (0.006) | 0.104 (0.010) | **$8.24 \times 10^{-6}$** | 0.88 (0.08, $1.38 \times 10^{-1}$) |
| PCA | 0.038 (0.007) | 0.055 (0.007) | $8.59 \times 10^{-2}$ | 0.83 (0.14, $2.13 \times 10^{-1}$) | MCH | 0.114 (0.017) | 0.224 (0.036) | **$5.73 \times 10^{-3}$** | 0.87 (0.12, $2.52 \times 10^{-1}$) |
| TL | 0.075 (0.023) | 0.062 (0.014) | $6.29 \times 10^{-1}$ | **0.61 (0.20, $4.63 \times 10^{-2}$)** | MCHC | 0.040 (0.007) | 0.083 (0.014) | **$6.01 \times 10^{-3}$** | 0.84 (0.15, $2.75 \times 10^{-1}$) |
| BMI | 0.141 (0.008) | 0.181 (0.007) | **$1.68 \times 10^{-4}$** | **0.84 (0.04, $1.53 \times 10^{-5}$)** | HGB | 0.050 (0.006) | 0.111 (0.013) | **$2.04 \times 10^{-5}$** | **0.79 (0.10, $3.70 \times 10^{-2}$)** |
| height | 0.327 (0.018) | 0.286 (0.015) | $8.01 \times 10^{-2}$ | **0.86 (0.04, $2.57 \times 10^{-4}$)** | MONO | 0.059 (0.010) | 0.163 (0.019) | **$1.27 \times 10^{-6}$** | **0.80 (0.09, $2.99 \times 10^{-2}$)** |
| DBP | 0.046 (0.005) | 0.106 (0.005) | **$2.16 \times 10^{-17}$** | **0.73 (0.06, $6.70 \times 10^{-6}$)** | NEUT | 0.087 (0.011) | 0.026 (0.005) | **$4.46 \times 10^{-7}$** | **0.77 (0.06, $2.42 \times 10^{-4}$)** |
| SBP | 0.056 (0.006) | 0.109 (0.004) | **$1.99 \times 10^{-13}$** | **0.71 (0.05, $5.62 \times 10^{-9}$)** | EO | 0.055 (0.011) | 0.130 (0.013) | **$1.06 \times 10^{-5}$** | **0.76 (0.09, $5.98 \times 10^{-3}$)** |
| PP | 0.037 (0.004) | 0.094 (0.004) | **$7.07 \times 10^{-24}$** | **0.73 (0.07, $8.80 \times 10^{-5}$)** | BASO | 0.039 (0.014) | 0.058 (0.007) | $2.25 \times 10^{-1}$ | **0.63 (0.12, $2.07 \times 10^{-3}$)** |
| HDL | 0.154 (0.033) | 0.241 (0.061) | $2.10 \times 10^{-1}$ | - | LYMPH | 0.057 (0.010) | 0.144 (0.011) | **$4.85 \times 10^{-9}$** | 0.84 (0.10, $8.26 \times 10^{-2}$) |
| LDL | 0.058 (0.012) | 0.171 (0.029) | **$3.18 \times 10^{-4}$** | 0.77 (0.18, $1.97 \times 10^{-1}$) | WBC | 0.070 (0.008) | 0.139 (0.013) | **$6.17 \times 10^{-6}$** | **0.75 (0.06, $3.85 \times 10^{-5}$)** |
| TC | 0.054 (0.009) | 0.183 (0.026) | **$2.75 \times 10^{-6}$** | 0.92 (0.11, $4.79 \times 10^{-1}$) | | | | | |

We first employed linkage disequilibrium score regression (LDSC) to estimate SNP-based heritability for every trait in each population [30]. Next, to evaluate genetic similarity and diversity of these traits across populations, we calculated the global trans-ethnic genetic correlation (*ρ_g_*) via popcorn [12]. We identified common trait-associated SNPs via the conditional false discovery rate (cFDR) and conjunction conditional false discovery rate (ccFDR) methods in the two populations [27, 28].

Relying on cFDR and ccFDR, for each trait we divided all analyzed SNPs into four incompatible groups. For every trait we estimated the marginal genetic correlation (*r_m_*) of SNP effect sizes in each of the four groups using MAGIC [25]. For common trait-associated SNPs, we examined the heterogeneity in genetic effect on the trait across EAS and EUR populations via Cochran’s Q test.

Then, we obtained the two LD scores and MAF for every SNP in the four groups based on genotypes available from EAS or EUR individuals in the 1000 Genomes Project, and calculated their coefficient of variation of LD scores (LDCV) and coefficient of variation of MAF (MAFCV) across the populations to investigate whether there exist different patterns of LD and MAF for trait-associated SNPs compared to those null ones.

Finally, to further demonstrate the direction of genetic differentiation, for each trait we conducted a GRS analysis [29]. We also investigated whether the sample size difference could influence our findings with regards to genetic similarity and diversity of traits between the EAS and EUR populations.

### Heritability and trans-ethnic genetic correlation

#### Estimated heritability

We first present the estimated SNP-based heritability (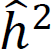) of these traits. It was shown some traits (e.g., height and schizophrenia [SCZ]) were more heritable, with a large heritability estimate; but other (e.g., ischemic stroke [IS] and atopic dermatitis [AD]) exhibited low heritability (**Table 1**). Although the estimates of heritability were highly correlated between the EAS and EUR populations (Pearson’s correlation=0.738, *P*=1.83×10 ^-7^), there still existed an obvious trans-ethnic distinction in heritability. For example, platelet count (PLT) showed the maximal difference in estimated heritability, with 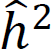=1.1% (se=1.4%) in EAS but 20.1% (se=1.8%) in the EUR population; in contrast, the heritability of atrial fibrillation (AF) was much larger in the EAS population compared to that in the EUR population (9.5% (se=2.7%) vs. 2.0% (se=0.3%)). By carrying out an approximate normal test, we observed that 70.3% (=26/37) of heritability estimates were significantly different across the two populations (FDR<0.05) (**Table 1**), largely reflecting diversity in polygenic genetic architecture across traits and ancestral groups.

We did not find a substantial correlation between sample size and heritability in both populations (Pearson’s correlation=0.112 with *P*=0.508 in the EAS population, and Pearson’s correlation=-0.025 with *P*=0.884 in the EUR population). However, as expected, we indeed discovered suggestive evidence that larger sample size can lead to more accurate estimate of heritability, with Pearson’s correlation=-0.266 in the EAS population and -0.102 in the EUR population between sample size and standard error of heritability. We further used the coefficient of variation of sample sizes (NCV) to measure the difference of sample sizes, but did not observe a significant correlation between NCV and the coefficient of variation of heritability (Pearson’s correlation=0.029, *P*=0.867).

#### Estimated trans-ethnic genetic correlation

The trans-ethnic genetic correlation estimates of six traits (i.e., AF, AD, breast cancer [BRC], high-density lipoprotein cholesterol [HDL], age at menarche [AAM], and triglyceride [TG]) were larger than one and thus not included in the following descriptions. All the traits exhibited positive trans-ethnic genetic correlation between the EAS and EUR populations (**Table 1**), with 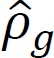 ranging from 0.53 (se=0.11) for adult-onset asthma (AOA) to 0.98 (se=0.17) for hemoglobin A1c (HbA1c).

All the trans-ethnic genetic correlation estimates were significantly larger than zero (*H*_0_: *ρ_g_*=0), but 61.3% (=19/31) were less than one (*H*_0_: *ρ_g_*=1) (FDR<0.05), indicating potential heterogeneity in genetic effects across populations. The average of 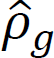 across traits was 0.79, and the average of 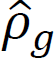 for binary phenotypes was 0.74 (se=0.15), which was slightly (although not significantly, *P*=0.217, possibly due to limited number of binary phenotypes under analysis) smaller than that for continuous ones (the average of 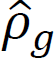=0.81 (se=0.09)).

#### Comparison for trans-ethnic genetic correlation, cross-trait trans-ethnic genetic correlation, and cross-trait genetic correlation

Furthermore, as an empirical comparison, we calculated cross-trait trans-ethnic genetic correlation and anticipated that the cross-trait correlation should be on average much smaller than that for the same traits in the two populations because of greatly distinct genetic foundations. As expected, it was found the same traits generally showed much greater genetic similarity compared to distinct traits although some of them, such as three lipids including low-density lipoprotein cholesterol [LDL], total cholesterol [TC], and TG, as well as three blood pressures including diastolic blood pressure [DBP], systolic blood pressure [SBP], and pulse pressure [PP], also exhibited relatively high cross-trait trans-ethnic genetic correlation (**Figure 1A** and **Table S1**). For instance, the cross-trait trans-ethnic genetic correlation ranged from -0.62 to 0.79, approximately 47.0% were negative, with an average of only 0.02 (**Figure 1B**).

**Figure 1.**
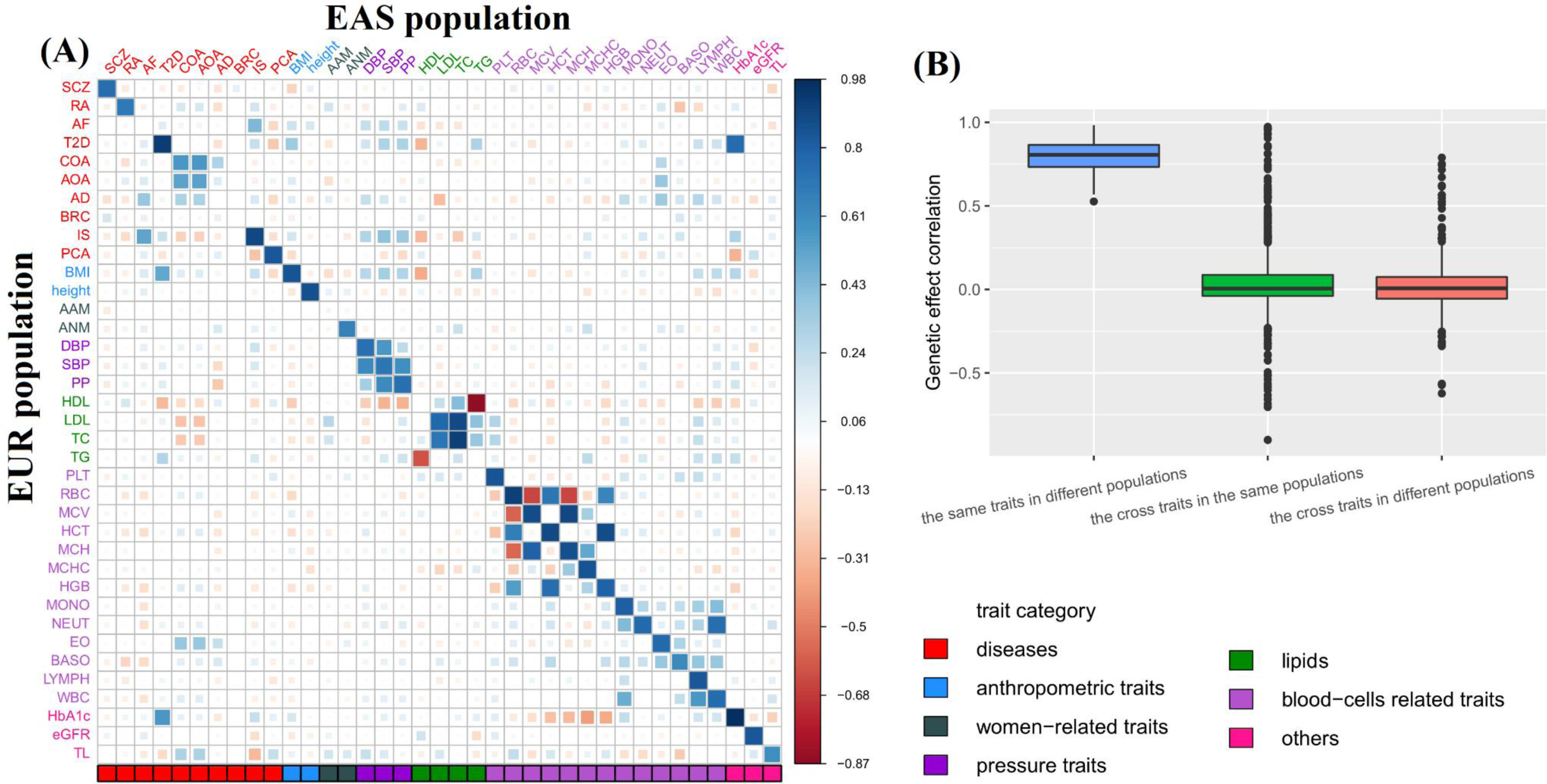
(**A**) Estimated trans-ethnic genetic correlation of 37 traits between the EAS and EUR populations. Elements in the diagonal represent e trans-ethnic genetic correlation for the same traits in the two populations, while elements in the off-diagonal represent the trans-ethnic enetic correlation for two different traits in the two populations. (**B**) Comparison of the estimated genetic correlation for the trait in the same opulation, two different traits in the same population, and two different traits in the EAS and EUR populations.

We also included the cross-trait genetic correlation in the same population as a reference. Totally, it was seen that the cross-trait genetic correlations in the same population were comparable to the cross-trait trans-ethnic genetic correlations across different populations but were much lower than the trans-ethnic genetic correlation of the same trait in diverse populations (**Figure 1B** and **Table S2**).

We did not discover a substantial connection between NCV and 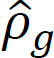 (Pearson’s correlation=-0.148, *P*=0.426), implying that the imbalanced sample sizes would not significantly affect the estimate of trans-ethnic genetic correlation. However, we observed that the difference of sample sizes in a pair of traits could affect the significance of 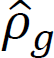. More specifically, 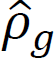 trended to be statistically different from one if larger imbalance of sample sizes was present (NCV=0.43 (se=0.25) for the 17 non-significant traits vs. 0.69 (se=0.27) for the 14 significant ones, *P*=0.010 in terms of the approximate normal test).

#### Associated SNPs for each trait in the EAS or EUR population

To discover trait-associated SNPs shared across the EAS and EUR populations, we carried out the cFDR analysis using a set of distinct SNPs [27, 28], with the number of significant associations displayed in **Table 2**. In general, more associated SNPs (cFDR<0.05) were discovered for most of the analyzed traits (94.6%=35/37, except for telomere length [TL] and HbA1c) in the EUR population (denoted by *f*_01_*+f*_11_ in **Table 2**) compared to the EAS population (denoted by *f*_10_*+f*_11_ in **Table 2**). This is a direct consequence of higher power due to larger sample sizes of EUR traits (**Figure 2A-B**) and implies that additional trait-associated loci would be detected if increasing samples especially in the EAS population. Only a few of SNPs were detected for some traits such as AD and IS, but much more associated SNPs were identified for others (e.g., BMI and height), partly indicating the polygenic nature of these traits [3, 31].

**Table 2.** Number of associated SNPs (cFDR/ccFDR<0.05) for traits in the EAS and EUR populations

| trait | $k$ | $f_{10}$ | $f_{01}$ | $f_{11}$ | $\hat{\pi}_{11}(\%)$ | $P_{LRT}$ | trait | $k$ | $f_{10}$ | $f_{01}$ | $f_{11}$ | $\hat{\pi}_{11}(\%)$ | $P_{LRT}$ |
| --- | --- | --- | --- | --- | --- | --- | --- | --- | --- | --- | --- | --- | --- |
| SCZ | 458,236 | 154 | 4,295 | 605 | 12.0 | $1.8 \times 10^{-266}$ | TG | 121,414 | 162 | 259 | 181 | 30.1 | $7.3 \times 10^{-86}$ |
| RA | 203,961 | 450 | 781 | 881 | 41.7 | $1.8 \times 10^{-266}$ | HbA1c | 124,962 | 88 | 50 | 106 | 43.4 | $3.3 \times 10^{-58}$ |
| AF | 236,293 | 166 | 1,262 | 242 | 14.5 | $1.2 \times 10^{-51}$ | eGFR | 236,693 | 805 | 3,413 | 933 | 18.1 | $1.8 \times 10^{-266}$ |
| T2D | 475,656 | 2,897 | 4,331 | 1,904 | 20.8 | $1.8 \times 10^{-266}$ | AAM | 319,439 | 35 | 6,271 | 340 | 5.1 | $7.6 \times 10^{-174}$ |
| COA | 250,069 | 259 | 1,877 | 421 | 16.5 | $6.2 \times 10^{-259}$ | ANM | 137,069 | 31 | 296 | 85 | 20.6 | $2.3 \times 10^{-48}$ |
| AOA | 249,871 | 239 | 970 | 432 | 26.3 | $1.1 \times 10^{-217}$ | PLT | 222,492 | 793 | 2,929 | 1,205 | 24.5 | $1.8 \times 10^{-266}$ |
| AD | 274,668 | 81 | 84 | 80 | 32.7 | $1.2 \times 10^{-51}$ | RBC | 222,589 | 413 | 2,258 | 890 | 25.0 | $1.8 \times 10^{-266}$ |
| BRC | 337,234 | 3 | 2,274 | 71 | 3.0 | $2.8 \times 10^{-24}$ | MCV | 222,587 | 790 | 2,597 | 1,273 | 27.3 | $1.8 \times 10^{-266}$ |
| IS | 246,666 | 10 | 30 | 39 | 49.4 | $1.6 \times 10^{-25}$ | HCT | 222,528 | 141 | 1,691 | 457 | 20.0 | $1.3 \times 10^{-256}$ |
| PCA | 247,201 | 134 | 641 | 293 | 27.4 | $2.4 \times 10^{-171}$ | MCH | 222,441 | 661 | 2,308 | 1,184 | 28.5 | $1.8 \times 10^{-266}$ |
| TL | 125,043 | 23 | 17 | 43 | 51.8 | $9.1 \times 10^{-48}$ | MCHC | 222,580 | 233 | 817 | 338 | 24.4 | $2.4 \times 10^{-213}$ |
| BMI | 118,899 | 438 | 11,818 | 768 | 5.9 | $1.8 \times 10^{-134}$ | HGB | 222,563 | 132 | 1,725 | 339 | 15.4 | $1.9 \times 10^{-226}$ |
| height | 139,126 | 3,528 | 5,292 | 2,616 | 22.9 | $1.8 \times 10^{-266}$ | MONO | 222,622 | 147 | 2,524 | 440 | 14.1 | $3.8 \times 10^{-160}$ |
| DBP | 214,705 | 84 | 13,624 | 468 | 3.3 | $2.1 \times 10^{-128}$ | NEUT | 222,435 | 170 | 2,020 | 309 | 12.4 | $4.3 \times 10^{-201}$ |
| SBP | 213,840 | 165 | 13,874 | 642 | 4.4 | $6.1 \times 10^{-150}$ | EO | 222,395 | 114 | 2,190 | 334 | 12.7 | $6.9 \times 10^{-184}$ |
| PP | 213,862 | 46 | 11,069 | 409 | 3.5 | $2.7 \times 10^{-125}$ | BASO | 222,525 | 143 | 598 | 200 | 21.3 | $2.7 \times 10^{-90}$ |
| HDL | 122,159 | 218 | 305 | 227 | 30.3 | $1.2 \times 10^{-104}$ | LYMPH | 222,490 | 32 | 2,307 | 263 | 10.1 | $1.3 \times 10^{-160}$ |
| LDL | 121,447 | 94 | 287 | 178 | 31.8 | $1.0 \times 10^{-112}$ | WBC | 222,550 | 343 | 2,451 | 555 | 16.6 | $6.7 \times 10^{-272}$ |
| TC | 122,106 | 151 | 367 | 275 | 34.7 | $3.9 \times 10^{-170}$ | | | | | | | |

**Note:** $k$ is the total number of SNPs analyzed by cFDR; $f_{10}$ and $f_{01}$ are the number of identified SNPs that are only associated with the trait in the EAS or EUR population, respectively, and $f_{11}$ is the number of shared associated SNPs; $\hat{\pi}_{11}$ is the proportion of trait-associated SNPs that are shared by both populations estimated in terms of the four-group model; $P_{LRT}$ is the adjusted $P$ value of genetic overlap obtained by the LRT method in the four-group model.

**Figure 2.**
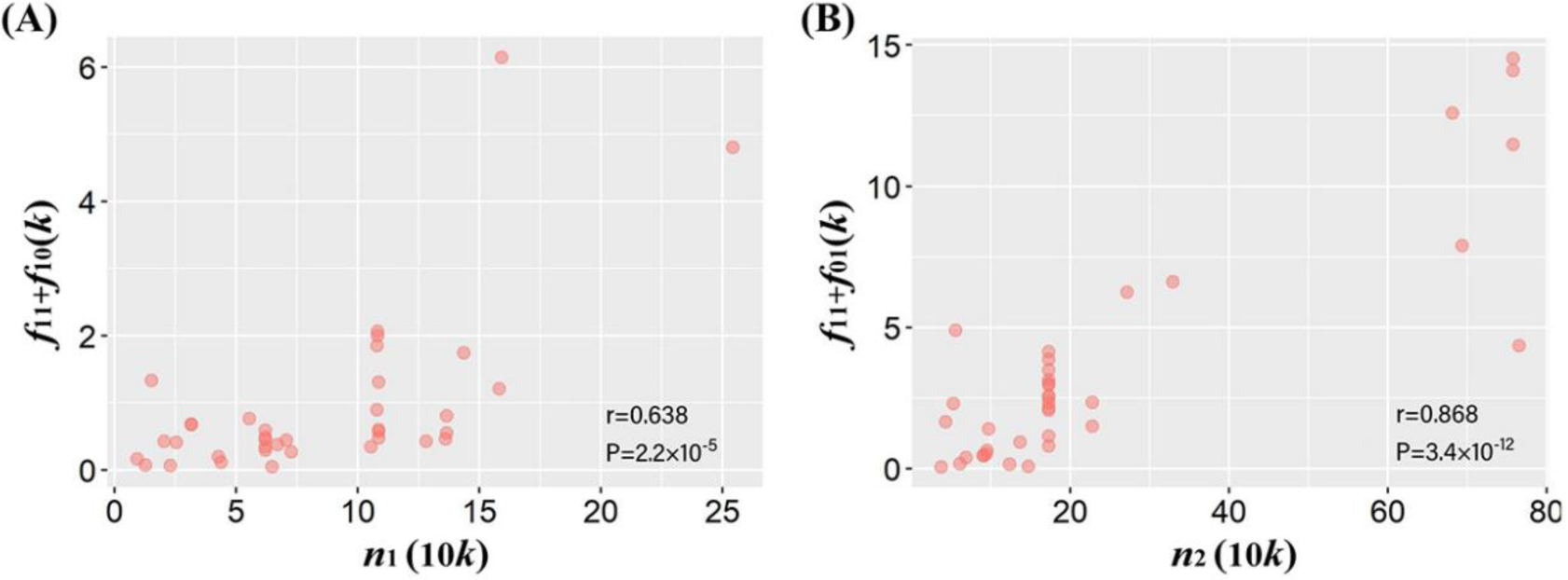
(**A**) Relationship between *n*_1_ and *f*_11_+*f*_10_; (**B**) Relationship between *n*_2_ and *f*_11_+*f*_01_. Here, *f*_10_ and *f*_01_ are the number of identified associated SNPs for the trait in the EAS or EUR population, respectively, and *f*_11_ is the number of shared trait-associated SNPs; *n*_1_ and *n*_2_ stand for the effective sample size for traits in the EAS or EUR population; *r* stands for the Pearson’s correlation, with *P* the corresponding *P* value; *k* on the x-axis means 1,000 for identified trait-associated SNPs and 10*k* on the y-axis means 10,000 for sample size of traits.

#### Shared associated SNPs of traits between the EAS and EUR populations

Furthermore, we identified many significant SNPs shared across the EAS and EUR populations (ccFDR<0.05, denoted by *f*_11_ in **Table 2**). On average, 21.7% of associated SNPs were discovered simultaneously for the same traits in both populations. The LRT implemented under the four-group model framework also offered considerably strong evidence supporting common genetic foundation underlying each trait between the two population. The proportion of shared associated SNPs varied substantially across these traits, ranging from only 3.0% for BRC and 3.3% for DBP to 49.4% for IS and 51.8% for TL (**Table 2**).

On average, approximately 67.1% of trait-associated SNPs in the EUR population were also detected to be significant in the EAS population, but only 26.8% of trait-associated SNPs in the EAS population were replicated to be significant in the EUR population. Among these shared significant SNPs, 44.1% and 77.0% showed genome-wide significance (*P*<5×10 ^-8^) in the EAS or EUR population (**Table S3**). We observed that the number of population-common associated loci (i.e., *f*_11_) was negatively correlated with NCV (Pearson’s correlation=-0.095, with a marginally significant *P* value of 0.057), indicating that smaller difference of sample sizes in a pair of traits (e.g., increasing the sample size of traits in the EAS population) might lead to more discoveries of shared SNPs.

### Characteristics of shared associated SNPs

#### Similarity and heterogeneity of associated SNPs across populations

It needs to highlight that we can divide all analyzed SNPs into four groups based on the associations identified above: (i) null SNPs; (ii) EAS-specific associated SNPs; (iii) EUR-specific associated SNPs; (iv) population-common associated SNPs. For these SNPs, it is shown that population-common associated SNPs often exhibited a maximal positive correlation in effect sizes compared to population-specific associated SNPs and null ones (**Figure 3A** and **Table S4**). For example, the marginal genetic correlation of effect sizes for shared trait-associated SNPs was 0.92 (se=0.04) for TL, followed by 0.81 (se=0.02) for white blood cell count (WBC) and 0.80 (se=0.03) for NEUT, with an average estimate of 0.61 (se=0.04) across these traits, which was much higher than that for EAS-specific (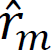 =0.29 (se=0.05)) or EUR-specific (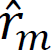 = 0.33 (se=0.05)) trait-associated SNPs or null SNPs (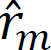 =0.09 (se=0.02)).

**Figure 3.**
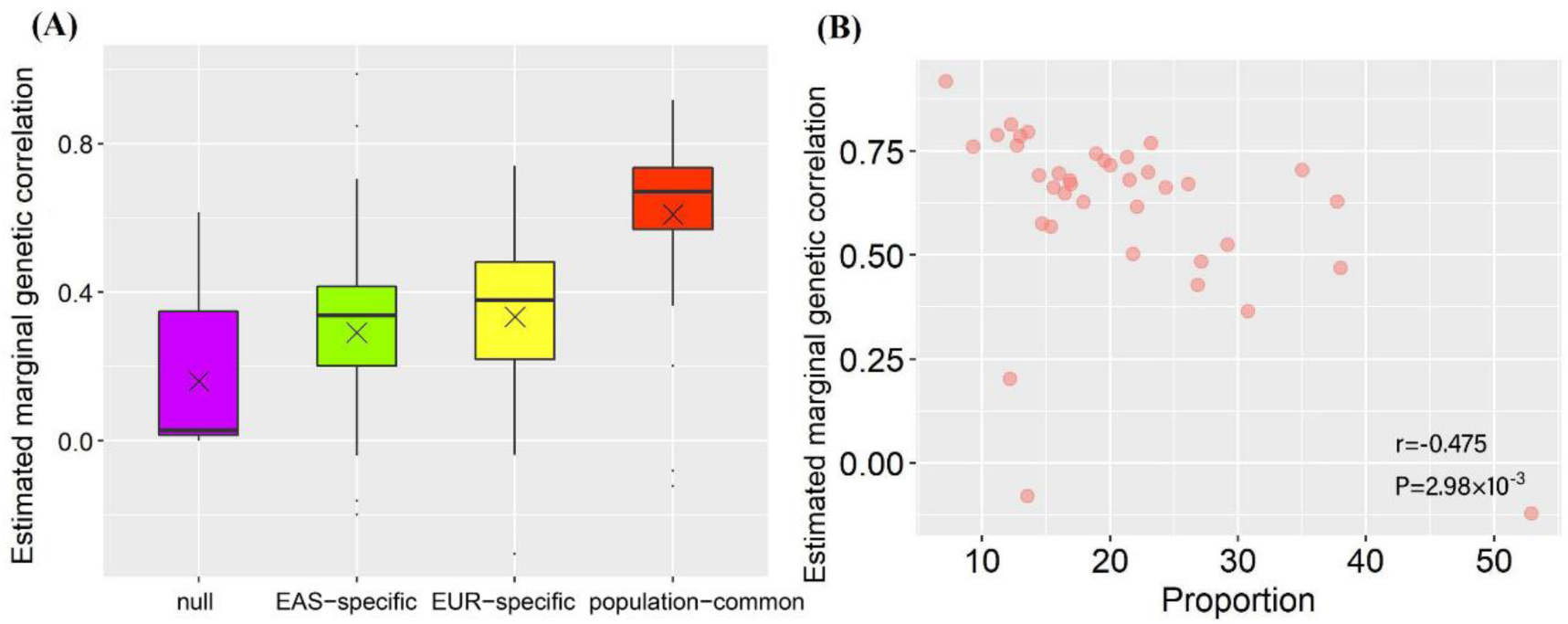
(**A**) Estimated marginal genetic correlation of effect sizes for SNPs in the four incompatible groups; (**B**) Relationship between proportions of genetic effect heterogeneity of shared associated SNPs and the cross-population marginal genetic correlations of SNP effect sizes; × means the median.

We found that the effect size slopes of population-common associated SNPs of eight traits (i.e., AF, AD, BRC, TL, SBP, PP, age at natural non-surgical menopause [ANM], and basophil count [BASO]) were not significantly different from one (FDR>0.05) (**Supplement File** and **Figure S1**), suggesting that effect sizes of shared associated SNPs are considerably consistent in magnitude for these traits; whereas great deviations of the estimated slopes from one were observed for the remaining 29 traits (FDR<0.05) (e.g., HbA1c, T2D, estimated glomerular filtration rate (eGFR), SCZ and AAM), indicating substantial trans-ethnic diversity of SNP effect sizes for these traits. In addition, we did not detect substantial evidence supporting the influence of imbalanced sample sizes (i.e., NCV) on the estimated slope (*P*=0.053).

We next performed a heterogeneity test using Cochran’s Q test and again focused only on population-common associated SNPs because the population-specific associated SNPs can be naturally considered to be heterogeneous (i.e., theoretically, their SNP effect sizes were non-zero in one population but zero in another population). On average, approximately 20.8% of the common trait-associated SNPs, ranging from 7.1% for TL to 52.9% for AAM, showed heterogeneous genetic effect on traits between the two ancestral populations after Bonferroni’s correction for multiple comparisons (**Table S5**). We here used Bonferroni’s method to take the LD among local SNPs into account as it was much more stringent compared to FDR. The high heterogeneity in SNP effect sizes for AAM was in agreement with a prior finding that AAM-associated SNPs often exhibited distinct effect sizes across populations [32]. It can be expected that greater proportion of shared trait-associated SNPs having heterogeneous effects would lead to weaker trans-ethic marginal genetic correlation (Pearson’s correlation=-0.48, *P*=2.98×10 ^-3^) (**Figure 3B**). However, we found little evidence supporting the influence of the sample size difference in a pair of traits (i.e., NCV) on the proportion of population-common SNPs with heterogeneous effects (*P*=0.771).

#### Difference in LD, MAF, and F_st_ for trait-associated SNPs across populations

First, we observed that, except for three lipid traits (i.e., TL, HDL, and LDL), all the remaining traits showed substantial different variations in LD for SNPs in various groups between the EAS and EUR populations (FDR<0.05) (**Figure 4A** and **Figure S2**). The average coefficient of variation of LD scores (LDCV) for population-common SNPs was smaller compared to that for null SNPs (0.22 (se=0.02) vs. 0.30 (se=0.02), *P*=1.04×10 ^-17^), for these population-specific loci in EAS (0.22 (se=0.02) vs. 0.28 (se=0.04), *P*=1.82×10 ^-9^) or EUR (0.22 (se=0.02) vs. 0.28 (se=0.03), *P*=1.52×10^-1^).

**Figure 4.**
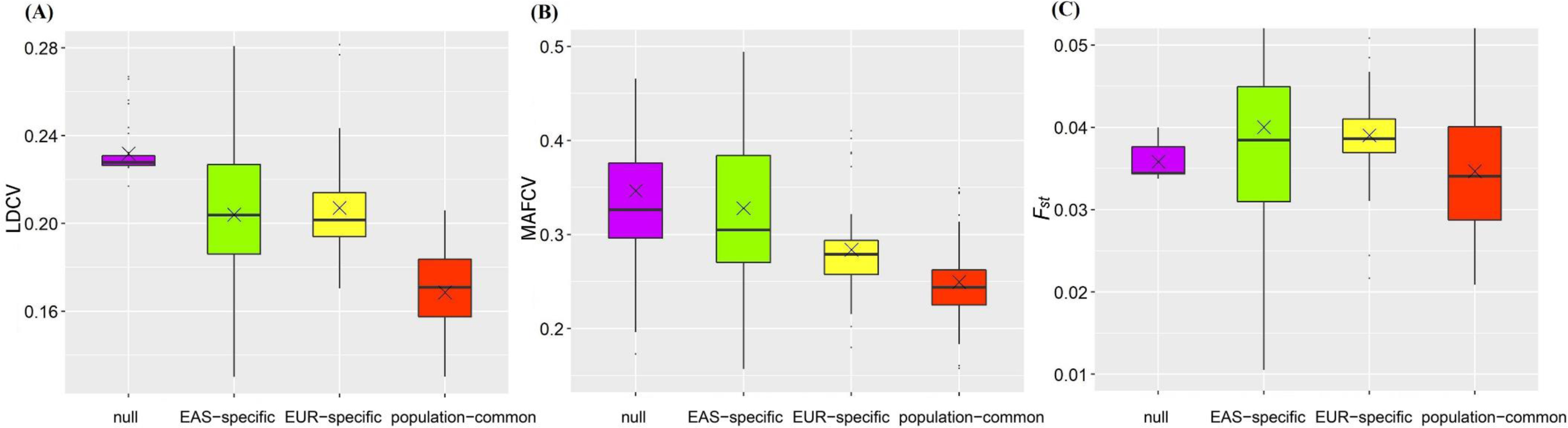
(**A**) Distribution for the average LDCV across all analyzed traits in the four groups of SNPs; (**B**) Distribution for the average MAFCV cross all analyzed traits in the four groups of SNPs; (**C**) Distribution for the average *F_st_* across all analyzed traits in the four groups of SNPs; × eans the median.

On the other hand, we observed that, except for three traits (i.e., HDL, TG, and ANM), all the remaining traits showed substantially different MAFCV for SNPs in distinct groups between the EAS and EUR populations (FDR<0.05) (**Figure 4B** and **Figure S3**). On average, the MAFCV for population-common loci were much smaller compared to that for null SNPs (0.33 (se=0.07 vs. 0.46 (se=0.10), *P*=1.54×10 ^-12^), for these population-specific loci in EAS (0.33 (se=0.07) vs. 0.43 (se=0.11), *P*=7.30×10 ^-8^) or EUR (0.33 (se=0.07) vs. 0.40 (se=0.07), *P*=2.22×10 ^-8^).

We found that SNPs suffered from natural selection for more than half of traits (62.2%=23/37) (FDR<0.05) (**Figure S4**). For all analyzed traits, we observed that population-common loci tended to have a smaller mean *F_st_* compared with population-specific associated SNPs in EAS (0.056 (se=0.01) vs. 0.062 (se=0.02), *P*=0.032) or EUR (0.056 (se=0.01) vs. 0.063 (se=0.006), *P*=6.77×10 ^-4^), and they also showed a lower mean *F_st_* relative to null ones (0.056 (se=0.01) vs. 0.060 (se=0.001), with a marginally significant *P*=0.086) (**Figure 4C**). Furthermore, we did not observe significant correlation between *F_st_* and LDCV (*P*=0.781) as well as between *F_st_* and MAFCV (*P*=0.602) (**Figure S5**), indicating the between-population diversity of LD and MAF patterns is not possibly confounded by the differentiation in *F_st_*.

#### Overall and partial GRS of trait-associated SNPs

First, we considered the GRS calculated from all associated SNPs (i.e., *f*_10_+*f*_11_ for EAS or *f*_01_+*f*_11_ for EUR) as an overall measurement of genetic effect on a given trait in each population. Among these, most of the traits had a substantially different GRS between the two populations (**Figure S6**). For example, six traits (i.e., RA, T2D, childhood-onset asthma [COA], AOA, BRC and prostate cancer [PCA]) showed a higher overall genetic effect on EAS, while some (i.e., SCZ, AF, AD and IS) displayed a larger overall genetic effect on EUR.

We further generated the GRS computed with only population-common associated SNPs (i.e., *f*_11_) or only population-specific associated SNPs (i.e., *f*_10_ for EAS or *f*_01_ for EUR), respectively. We viewed these new GRSs to quantify a measurement of partial genetic effect because only part of associated SNPs is employed. Interestingly, 16 traits (i.e., RA, T2D, AD, PCA, DBP, SBP, LDL, TC, TG, eGFR, ANM, mean corpuscular hemoglobin [MCV], hematocrit [HCT], monocyte count [MONO], eosinophil count [EO] and WBC) had consistent partial genetic effect compared to the overall one. Specifically, 15 traits (e.g., AF, COA, AOA, BRC, BMI, height, PP, HDL, HbA1c, RBC, mean corpuscular hemoglobin [MCH], mean corpuscular hemoglobin concentration [MCHC], hemoglobin concentration [HGB], NEUT, and BASO) showed consistent partial genetic effect in terms of GRS calculated with EAS- or EUR-specific associated SNPs, whereas only 4 traits (i.e., SCZ, AAM, PLT, and lymphocyte count [LYMPH]) exhibited consistent partial genetic effect in terms of GRS calculated with shared associated SNPs. Particularly, IS and TL exhibited a completely opposite partial genetic effect in terms of the EAS/EUR-specific associated SNPs or population-common associated SNPs compared with the overall genetic impact measured with all trait-associated SNPs.

### Discussion

#### Summary of results in the present study

The present study has analyzed a total of 37 complex traits and sought to compare shared and distinct genetic components underlying them between the EAS and EUR populations. We discovered there existed pervasive consistence in heritability and trans-ethnic genetic correlation for these traits. Additionally, it needs to highlight that the trans-ethnic genetic correlation of continuous traits was on average higher compared to those binary traits, which may be due to the loss of information when converting some continuous phenotypes into binary ones (e.g., using HbA1c to define T2D) or in part reflects the discrepancy of classification and diagnosis of diseases in distinct populations.

Using cFDR [27, 28] we detected a great number of population-common association signals as well as many population-specific associated SNPs. A further exploration demonstrated these shared trait-associated SNPs generally showed the maximal positive correlation in effect sizes compared to population-specific trait-associated SNPs and null ones [7, 25]. Interestingly, we observed that even the shared association signals also exhibited a considerable degree of heterogeneity in genetic influence on traits across the EAS and EUR populations.

Furthermore, we revealed that population-specific associated SNPs were often more possible to suffer from natural selection compared with population-common associations, whereas population-common associated SNPs often displayed more consistent patterns in LD and MAF across continental populations.

Especially, the GRS analysis showed that population-common and population-specific associated SNPs have potentially different genetic influence on phenotypic variation and that the genetic differentiation from associated SNPs may at least partly explain the observed phenotypic variation across diverse ancestral groups. For example, the average GRS of SCZ in the EUR population was on average higher than that in the EAS population, partly contributing to the observation that SCZ was more prevalent in individuals of EUR ancestry than those of EAS ancestry [33]. The mean GRS of T2D was higher in the EAS population than that in the EUR population, supported by the observed incidence difference between the populations [34, 35]. It was reported that the absolute risk of T2D tended to be higher among Asians compared with Caucasians for any given level of body mass index [BMI] and waist-hip ratio [34]. As another example, for PCA higher mean GRS was observed in EAS compared to EUR, in line with a previous study which indicated that more than half of SNPs showed larger effect sizes in EAS than EUR [36].

However, we also observed patterns that seemed to be opposite against prior findings. For example, it was shown the mean GRS of AD in EUR was higher than that in EAS, in contrast to previous observation that Asians and Pacific Islanders were seven-fold more likely than whites to be diagnosed with AD because stronger Th17/Th22 polarization and mutations in immune-related genes such as *DEFB1* [37–39]. These inconsistent results can be expected because of the complicated nature of these traits, and can be explained by gene-gene/gene-environment interaction, ethnic difference and genetic factors that are largely underappreciated in our current study.

#### Comparison our discoveries to prior studies

Like the findings obtained from prior studies that however often focused only on a single trait [11, 40, 41], our work, which considered much more traits and diseases, further confirmed extensive genetic overlap and identified a large number of common associated genetic loci across different populations. From a biological perspective, there is no doubt about the widespread existence of population-shared risk variants because the risk variants targeted by GWASs are often common genetic loci that are believed to be of ancient origin and largely shared among different populations.

As revealed in our study, while some of associated SNPs vary substantially across populations, common associated SNPs in the EAS and EUR populations nevertheless often show much more similar effect sizes and effect directions; therefore, at least part of trait-associated SNP mapping results discovered in one population can be transferred to the other populations [42]. It needs to highlight that we may underestimate the degree of genetic sharing between various ancestry groups because the much smaller number of individuals in the EAS GWASs reduces power to detect homogeneity of effect compared to the EUR GWASs. These findings are largely consistent with some recent discoveries that most common causal SNPs were shared across the EAS and EUR populations, high-posterior SNPs identified by fine-mapping often showed highly correlated effects, and population-specific genetic regions likely harbored common trait-associated SNPs which however failed to be detected in the other GWASs due to differences in LD, MAF, and/or sample size [43].

On the other hand, despite highly shared genetic architecture, we indeed found evidence of heterogeneity at trait-associated variants, which meanwhile challenges in assessing the transferability of risk variants across different ethnic populations based on associations discovered in EUR GWASs [44, 45]. For example, although the trans-ethnic marginal genetic correlation for population-common associated SNPs of HbA1c between EAS and EUR was as high as 0.63, heterogeneity was detected at 37.7% shared associated loci. This diversity may point to the difference in clinical definitions and phenotype measurements [46]; and it can be in part explained by interaction between gene-gene and gene-environment [47].

The genetic difference may also underlie the well-known trans-ethnic dissimilarities in prevalence or characteristics of the traits [48–52]. In our analysis, these population-specific association signals largely indicate the significant trans-ethnic difference, which exist in distinct LD and allele frequency [7, 53–55]. In addition, many studies have revealed that, unlike in most European ancestry populations, the population genetic history of non-European ancestry groups has undergone selective pressure due to the effects of malaria and other infectious diseases on erythrocytes [40, 56]. Another possible factor for the genetic inconsistency of complex traits between ancestry groups may be due to sample size difference and thus different statistical power between EAS and EUR studies.

#### Important scientific implications of our findings

Our findings regarding the degree of common or diverse genetic components of the traits across ancestral groups have important implication in practice. For example, theoretically, genetic correlation offers the maximal boundary of trans-ethnic genetic prediction power [5, 12]; however, both overall trans-ethnic correlation and marginal trans-ethnic correlation imply low accuracy when implementing genetic prediction in one population of interest on the basis of associated loci discovered in other populations [25], indicating the need to carry out GWASs with more ancestral groups.

In addition, it is helpful for aggregating multiple study cohorts across ethnicities to conduct trans-ethnic GWAS analysis [57–60], developing trans-ethnic genetic risk prediction [25, 61], and fine-mapping causal genetic variants in minority populations [62, 63]. It also holds the key to benefit more ethnic groups from current medical genomics researches [64–66]. All of these offer promising avenues in post-GWAS era by integrating trans-ethnic information.

#### Potential limitations

The present study is not without limitation. First, as mentioned before, the studies in EAS are in generally underpowered due to much smaller sample size compared to that in EUR and hence our results may be affected by power issues. The small sample size may also lead to unstable effect estimation for these SNPs. Moreover, as shown before, we cannot completely rule out the possibility that the imbalanced sample sizes can also partly interpret the observed trans-ethnic genetic difference in traits. For example, we found that sample size was significantly positively correlated with the number of trait-associated loci in both populations, and that the estimated trans-ethnic genetic correlation would become less significantly different from one when the difference of sample sizes in a pair of traits reduced. Thus, the external validation of our results with larger sample size especially for EAS GWASs is warranted.

Second, besides the difference in sample sizes, other discrepancies in study designs such as phenotypic definition, statistical methods, and covariate adjustment can be also contributable to the observed trans-ethnic genetic similarity and diversity. Examining and quantifying the relative contributions are imperative for understanding genetic heterogeneity across populations. However, compared to the difference in sample sizes which are already reported in summary statistics, the design discrepancies in GWAS are difficult to handle with only summary statistics. It needs large-scale individual-level data of phenotypes and genotypes, and is thus challenged by privacy concerns when sharing data [67].

Third, individuals of EAS and EUR ancestries are to a great extent genetically similar, whereas more major genetic differences are expected to be found between AFR and non-AFR populations [68]. Therefore, it is not clear whether our findings can be generalized to comparison in other populations such as EUR and AFR. Unfortunately, the number of GWASs performed in individuals of AFR descent is still too limited to enable comparative studies.

Fourth, our analysis only considered common SNPs (MAF>0.01) and ignored rare variants, which usually have a recent origin compared with common ones from ancient origin. Theoretically, rare risk variants might be more likely to be population-specific and could possibly carry a greater risk effect, which probably leads us to underestimate the genetic heterogeneity across populations.

Fifth, to our knowledge, this is the first time that the cFDR method has been employed in detecting trans-ethnic genetic overlap for a large range of complex traits. However, it implements association mapping at a fixed FDR level rather than the standard error measure such as family-wise error rate (FWER) or type I error rate which is more widely-used in a typical GWAS. FDR is more liberal compared to FWER; thus, we can discover a much larger number of trait-associated SNPs. Although the cFDR method has been well-established under the context of pleiotropy-informed association mapping in ancestry-matched populations [27, 28] and also demonstrated to be well-calibrated in our study (**Figures S7-S8**), its ability of controlling FWER is less understood. Consequently, we just considered cFDR as a powerful tool for discovering evidence of trans-ethnic genetic overlap in our application, and by no means attempted to replace the standard GWAS analysis strategy with cFDR nor the cFDR level (e.g., 0.05 used here) with the genome-wide significance level (e.g., 5×10 ^-8^).

#### Conclusions

Our work provides an in-depth understanding of similarity and diversity regarding genetic architecture for complex traits across diverse populations, and can assist in trans-ethnic association analysis, genetic risk prediction, and causal variant fine mapping.

### Materials and Methods

#### Summary statistics of 37 complex traits

We obtained summary statistics (e.g., marginal effect size and standard error) of 37 complex traits (10 binary and 27 continuous) analyzed on EAS-only or EUR-only individuals (**Table 1** and **Tables S6-S7**). These traits included lipids (e.g., TG), blood cell phenotypes (e.g., neutrophil [NEUT] and MONO), diseases (e.g., BRC, T2D, and prostate cancer [PCA]), and anthropometric phenotypes (e.g., BMI and height).

For each analyzed trait, we carried out stringent quality control in both populations by following previous work [12, 30, 69]: (i) filtered out SNPs without rs label; (ii) deleted non-biallelic SNPs and those with strand-ambiguous alleles; (iii) removed SNPs whose alleles did not match with those in the 1000 Genomes Project; (iv) excluded duplicated SNPs; (v) filtered out palindromic SNPs containing the same bases; (vi) kept only common SNPs (MAF>0.01 in each population) which were also included within the 1000 Genomes Project and shared between the EAS and EUR GWASs.

Here, MAF was calculated with genotypes of EAS (*N*=504) or EUR (*N*=503) individuals in the 1000 Genomes Project if missing in the original summary statistics data; the threshold value of 0.01 for MAF was selected as it was widely used in summary statistics-based studies [12, 30, 69]. We further aligned the effect allele of all remaining SNPs for each trait between the two populations.

#### Estimation of heritability and cross-trait genetic correlation in the same population

We first employed LDSC to estimate SNP-based heritability for each trait separately in the two populations [30]. The LDSC software (version v1.0.1) was downloaded from https://github.com/bulik/ldsc and the analysis was conducted with default parameter settings. The LD score was calculated with genotypes of SNPs (MAF>0.01 and the *P* value of Hardy Weinberg equilibrium test>1.0×10 ^-5^) with a 10Mb window on EAS or EUR individuals in the 1000 Genomes Project. Then, the LD score of SNP was regressed on the square of *Z*-statistic of the analyzed trait, with the regression slope offering an unbiased estimate for heritability. Besides quality control procedures described above, we here further removed SNPs located within the major histocompatibility complex region (chr6: 28.5∼33.5Mb) because of its complicated structure which was often difficult to estimate accurately from an external reference panel [30, 70]. Relying only on summary statistics and LD scores, LDSC can be also applied to calculate the cross-trait genetic correlation in the same population [69].

#### Estimation of trans-ethnic genetic correlation across populations

To evaluate genetic similarity and diversity for these traits across populations, we calculate the global trans-ethnic genetic correlation (*ρ_g_*) via popcorn [12]. Conceptually, *ρ_g_* is defined as the correlation between SNP effect sizes of the trait in various ancestral groups to measure the extent to which the same SNP exhibits the same or similar impact on phenotypic variation [12, 25, 26]. Methodologically, popcorn was proposed from the Bayesian perspective by assuming effect sizes of SNPs following an infinitesimal model [71], and can be considered as a natural trans-ethnic extension of LDSC. The trans-ethnic LD score of each SNP was downloaded from https://github.com/brielin/popcorn, which was calculated with genotypes of EAS or EUR individuals in the 1000 Genomes Project between the focal SNP and all the flanking ones within a 10Mb window. To obtain an unbiased estimate of trans-ethnic genetic correlation, we implement an unbounded estimation algorithm in popcorn, which likely leads to an estimate less than -1 or greater than 1.

### Identification of associated SNPs and shared genetic overlap across populations

#### Conditional false discovery rate and conjunction conditional false discovery rate

From a statistical perspective, we observe that the identification of trans-ethnic genetic overlap can be handled by applying the similar principle of detecting pleiotropic associations for genetically correlated traits in a single ancestral group. Over the past few years many methods have been developed for detecting pleiotropy [72–74]. Among those, cFDR is a novel pleiotropy-informed method to discover genetic overlap and can be viewed a novel extension of the classical FDR for a single trait in one population to the same trait in trans-ethnic cases [27, 28]. By integrating association results from multiple traits, this method could offer important sights into trans-ethnic genetic overlap and increased power to identify less significant association signals.

In our application context, the null hypothesis of FDR is no association between a SNP and the trait of focus in one population. Based on this definition and the principle of FDR, cFDR is logically defined as the posterior probability that a random SNP is null for the trait in one population given that the observed *P* values for the trait in both populations are less than a predetermined threshold. Formally, with two sets of *P* values as input, cFDR is calculated as 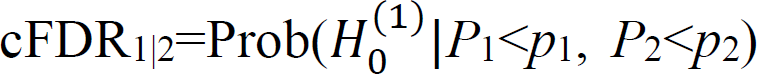, where *p*_1_ and *p*_2_ are the observed *P* values of a particular SNP for the trait in the two populations, respectively; and 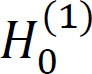 denotes the null hypothesis indicating there does not exist an association between the SNP and the trait in the first population. cFDR is efficiently estimated with an empirical Bayesian manner that was proposed for computing the local FDR [75].

As the principal and conditional positions for the trait in cFDR described above are exchangeable between the populations, cFDR_2|1_ is defined in a similar way. Therefore, ccFDR for identifying shared SNPs is simply expressed as ccFDR_1|2_=max(cFDR_1|2_, cFDR_2|1_), which is defined as the probability that a particular SNP has a false positive association with the trait in the two populations given the observed *P* values. Finally, SNPs with ccFDR less than a given significance threshold can be prioritized to be population-common SNPs. Although the traditional meta-analysis is also often applied in trans-ethic association studies [76–81]; however, the association discovered by meta-analysis cannot certainly suggest trans-ethic genetic overlap because such association might be only present in one special population.

Because cFDR and ccFDR are constructed for relatively independent SNPs, to generate uncorrelated SNPs, following prior work [82] we applied the LD pruning (the width of SNP window=50 and *r*^2^=0.1) in PLINK with genotypes of EAS or EUR individuals separately in the 1000 Genomes Project as the reference panel for LD calculation, and then combined the two sets of SNPs available from both populations (**Table S6**). In addition, based on findings observed in other studies [73, 74], genetic/phenotypic correlation between traits can result in inflated test statistics. Therefore, to minimize false discovery in our cFDR analysis, we further generated uncorrelated *Z*-statistics for every trait across populations by multiplying them by the inverse of a correlation matrix, which can be easily calculated in terms of the two statistics of null SNPs (e.g., SNPs with *P*>10^-4^) [72, 83]. This decorrelation strategy maximizes the transformed test statistics and the original ones [84]; therefore, it has the minimal influence on association identification. These uncorrelated *Z*-statistics were ultimately transformed into two-sided *P* values for the cFDR analysis based on normal approximation.

Our simulation studies already demonstrated that the cFDR method could maintain well-calibrated control of FDR at the given level and behaved better when identifying population-common trait-associated SNPs compared to the naïve minimum *P*-value method (**Supplement notes** and **Figures S7-S8**).

#### Various types of SNPs and four-group model

Relying on cFDR and ccFDR, for each trait we could divide all the analyzed SNPs into four incompatible groups: (i) not associated with the trait in neither population (i.e., null SNPs); (ii) only associated with the trait in the EAS population but not in the EUR population (i.e., EAS-specific associated SNPs); (iii) only associated with the trait in the EUR population but not in the EAS population (i.e., EUR-specific associated SNPs); (iv) associated with the trait in both the two populations (i.e., population-common associated SNPs).

To measure the degree of genetic components shared by the same trait across the populations, we further applied the four-group model [85] which examined SNPs in the four groups mentioned above. This model aims to estimate the proportions of SNPs in each group, and employs the LRT method to assess the statistical significance for overall trans-ethnic genetic overlap. Statistically, the four-group model assumes that *P* values of null SNPs (not associated with the trait in neither populations) follow the uniform distribution and *P* values of non-null SNPs (associated with the trait at least in one population) follow the Beta distribution.

### Genetic correlation and heterogeneity of SNPs between the two populations

#### Marginal genetic correlation across populations

For every trait we calculated the marginal genetic correlation (*r_m_*) of SNP effect sizes in each of the four groups using a recently proposed method called MAGIC [25]. Compared to the traditional Pearson’s correlation, which often underestimates the marginal genetic correlation thus leads to the so-called correlation attenuation because of failing to take the estimation error of effect sizes into account [25, 86], MAGIC has the advantage of generating unbiased correlation estimation by accounting for the uncertainty under the framework of measurement error model [87].

#### Linear regression for SNP effect sizes

We also carried out a simple linear model without the intercept term for only population-common associated SNPs of each trait by regressing effect sizes of SNPs in one population on those in another population. The slope of the linear regression model provides an indicator for the relative magnitude of effect sizes for shared trait-associated SNPs between the two populations. For common associated SNPs, we examined the heterogeneity in genetic effect of SNPs on the trait across EAS and EUR populations via Cochran’s Q test in the R metafor package.

### Characteristics of associated SNPs between EAS and EUR populations

#### Patterns for LD, MAF, and Wright’s fixation index

After detecting common associations, we wondered whether there exist different patterns of LD and MAF for trait-associated SNPs compared to those null ones. To this aim, we first obtained the two LD scores for every SNP in the four groups based on genotypes available from EAS or EUR individuals in the 1000 Genomes Project, and then calculated their LDCV across the populations. Here, coefficient of variation, rather than variance, was utilized because SNPs with higher LD scores tended to have greater variation between populations [7, 24]. In a similar way, we calculated MAFCV for each SNP between the two populations to evaluate how MAF varies between populations.

We further evaluated whether an identified trait-associated SNP had been under natural selection. If this was the case, then a substantial between-population differentiation would be observed in the allele frequency [24, 88]. To this aim, we applied the Wright’s fixation index (*F_st_*) to evaluate such an allele frequency diversity across populations under natural selection [24, 88, 89], and calculated *F_st_* of each SNP in the four groups for every trait with genotypes available from EAS and EUR individuals in the 1000 Genomes Project using PLINK.

To examine LDCV, MAFCV, and *F_st_* in the four groups, we carried out the Kruskal test for each trait, with the resulting *P* values being FDR corrected to account for multiple testing using the Benjamini-Hochberg procedure. We also performed a paired test to compare the average of LDCV, MAFCV, and *F_st_* across the traits while simply ignoring the uncertainty of the estimated average in each group.

#### Partial and overall genetic risk score analysis

To further demonstrate the direction of genetic differentiation, for each trait we conducted a GRS analysis [29]. The calculated GRS in part measures the stratification of the whole population based on estimates of individual’s genetic susceptibility. In our analysis the GRS of a given individual was simply computed as 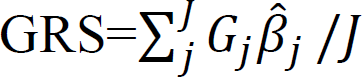, where *J* was the number of SNPs used and *G* represented the genotype (coded as 0, 1, or 2) available from the EAS or EUR individuals in the 1000 Genomes Project. For binary phenotypes, 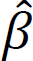 was the original marginal SNP effect size; whereas, for the compatibility across populations, for continuous phenotypes we re-scaled 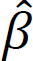 based on *Z*-statistic [90]; that is, 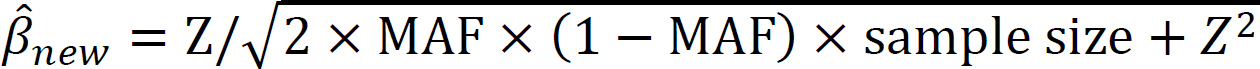.

For each pair of traits under analysis, three types of GRS were generated using three distinct sets of SNPs (**Table 2**), including population-specific loci (*J=f*_10_ or *f*_01_), population-common loci (*J=f*_11_) and population-associated loci (*J=f*_10_+*f*_11_ or *f*_01_+*f*_11_). For the convenience of description, we referred to the first two GRSs as partial GRS, while the third one as overall GRS.

### Exploring the influence of sample size difference

We finally attempted to investigate whether the sample size difference could influence our findings with regards to genetic similarity and diversity of traits between the EAS and EUR populations. First, after obtaining the heritability estimate and its standard error for a given trait in the two populations (denoted by 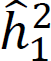 and 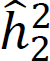, *se*_1_ and *se*_2_, respectively), we studied the influence of sample sizes of traits on the estimated heritability and its standard error.

Second, to assess the impact of sample size on the trans-ethnic diversity of estimated heritability, we employed NCV to measure the difference of sample sizes, and examined its relation with the coefficient of variation of heritability, the estimated slope of effect sizes for population-common SNPs and the proportion of population-common SNPs with heterogeneous effects.

Third, we implemented an approximation normal test to examine the significance of the difference in the estimated heritability between the two populations by calculating 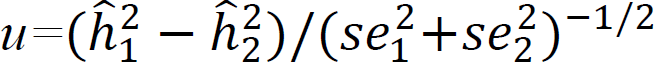. We obtained the *P* value of *u* using the standard normal distribution as the null distribution, with the issue of multiple testing corrected via by the Benjamini-Hochberg method. By doing this, we explicitly took the difference of sample sizes in traits into account by modeling its standard error when comparing heritability.

## Declarations

### Ethics approval and consent to participate

All methods were carried out in accordance with relevant guidelines and regulations (declaration of Helsinki).

### Consent for publication

Not applicable.

### Availability of data and materials

All data generated or analyzed during this study are included in this published article and its supplementary information file.

## Data Availability

All data produced in the present study are available upon reasonable request to the authors

## Acknowledgements

We thank all the GWAS consortia for making summary statistics publicly available for us and are grateful to all the investigators and participants contributed to those studies. The data analyses in the present study were carried out with the high-performance computing cluster that was supported by the special central finance project of local universities for Xuzhou Medical University.

## Authors’ contributions

PZ conceived the idea for the study. PZ and JZ obtained and cleared the datasets; TW, PZ, JQ, SZ and JZ performed the data analyses. TW, PZ, JQ, SZ, and JZ interpreted the results of the data analyses. TW, PZ and JZ wrote the manuscript with the help from other authors.

## Abbreviations

SNP: single-nucleotide polymorphism
GWAS: genome-wide association study
EUR: European
EAS: East Asian
LD: linkage disequilibrium
cFDR: conditional false discovery rate
ccFDR: conjunction conditional false discovery rate
MAF: minor allele frequency
GRS: genetic risk score
LDSC: linkage disequilibrium score regression
LDCV: coefficient of variation of
LD: scores
MAFCV: coefficient of variation of MAF
*F_st_*: Wright’s fixation index

