## Supplement File for "Similarity and diversity of genetic architecture for complex traits between East Asian and European populations"

### Supplementary File

### Supplement notes

We conducted a simple simulation study to assess the performance of conditional false discovery rate (cFDR) in identifying trans-ethnic genetic overlap. Because the cFDR method can be implemented with only summary statistics of the same trait available from two populations, we hence directly generated two sets of *P* values from a given bivariate normal (BVN) distribution under various scenarios.

Specifically, for the first set of *Z-*score in a pair, we drew them from BVN((0, 0), Σ) with a probability π_00_, or from BVN((*μ*_10_, 0), Σ) with a probability π_10_, or from BVN((0, 0), Σ) with a probability π_01_, or from BVN((*μ*_11_, 0), Σ) with a probability π_11_. For the second set of *Z-*score in a pair, we sampled them from BVN((0, 0), Σ) with a probability π_00_, or from BVN((0, 0), Σ) with a probability π_10_, or from BVN((0, *μ*_01_), Σ) with a probability π_01_, or from BVN((0, *μ*_11_), Σ) with a probability π_11_. We set Σ to be a two-dimensional identify matrix.

To speed up the computation, we set the number of independent SNPs under analysis to be 10000, 15000, or 20000, *μ*_10_=*μ*_01_=*μ*_11_=2, 3 or 4, and considered two settings of probabilities with π_11_≠0 (π_00_=0.40, π_10_=0.20, π_01_=0.20 and π_11_=0.20; andπ_00_=0.80, π_10_=0.05, π_01_=0.05 and π_11_=0.10) for evaluating the false discovery rate (FDR) and power. Note that, the magnitude of *μ*_10_ (or *μ*_01_ and *μ*_11_) quantified the strength of association, with larger value indicating stronger signal.

Afterwards, we transformed *Z-*statistic into *P* value based on the normal approximation. For both FDR control and power evaluation, we repeated the simulation in each setting 10^3^ times and displayed the average across these replicates.

Overall, it was found that cFDR can maintain correct FDR control and much powerful across our simulation scenarios compared to the minimum *P*-value method (minP), which was conservative and thus was less powered (**Figures S7-S8**).

### Supplement results

#### Table S3. Number of trait-associated SNPs (ccFDR<0.05) also showing genome-wide significance in the EAS and EUR populations

| trait | *G*_1_ | *G*_2_ | *P*_1_ (π_1_, %) | *P*_2_ (π_2_, %) | trait | *G*_1_ | *G*_2_ | *P*_1_ (π_1_, %) | *P*_2_ (π_2_, %) |
| --- | --- | --- | --- | --- | --- | --- | --- | --- | --- |
| SCZ | 62 | 495 | 50 (80.6) | 117 (23.6) | TG | 93 | 99 | 67 (72.0) | 74 (74.7) |
| RA | 403 | 672 | 304 (75.4) | 481 (71.6) | HbA1c | 24 | 25 | 20 (83.3) | 19 (76.0) |
| AF | 35 | 293 | 28 (80.0) | 102 (34.8) | eGFR | 117 | 601 | 85 (72.6) | 247 (41.1)) |
| T2D | 659 | 633 | 365 (55.4) | 382 (60.3) | AAM | 6 | 744 | 5 (83.3) | 155 (20.8) |
| COA | 99 | 545 | 47 (47.5) | 131 (24.0) | ANM | 21 | 53 | 13 (61.9) | 26 (49.1) |
| AOA | 93 | 259 | 82 (88.2) | 121 (46.7) | PLT | 248 | 694 | 192 (77.4) | 348 (50.1) |
| AD | 33 | 15 | 12 (36.4) | 9 (60.0) | RBC | 114 | 480 | 104 (91.2) | 283 (59.0) |
| BRC | 6 | 454 | 5 (83.3) | 50 (11.0) | MCV | 320 | 776 | 238 (74.4) | 457 (58.9) |
| IS | 3 | 4 | 3 (100.0) | 4 (100.0) | HCT | 27 | 330 | 24 (88.9) | 134 (40.6) |
| PCA | 66 | 157 | 54 (81.8) | 73 (46.5) | MCH | 307 | 759 | 226 (73.6) | 438 (57.7) |
| TL | 16 | 5 | 13 (81.3) | 3 (60.0) | MCHC | 92 | 315 | 64 (69.6) | 123 (39.0) |
| BMI | 55 | 1,309 | 45 (81.8) | 295 (22.5) | HGB | 19 | 366 | 16 (84.2) | 119 (32.5) |
| Height | 786 | 958 | 533 (67.8) | 619 (64.6) | MONO | 62 | 535 | 52 (83.9) | 213 (39.8) |
| DBP | 32 | 2,050 | 27 (84.4) | 252 (12.3) | NEUT | 61 | 351 | 40 (65.6) | 95 (27.1) |
| SBP | 62 | 1,940 | 58 (93.5) | 336 (17.3) | EO | 55 | 510 | 45 (81.8) | 141 (27.6) |
| PP | 30 | 1,605 | 28 (93.3) | 232 (14.5) | BASO | 61 | 163 | 42 (68.9) | 50 (30.7) |
| HDL | 122 | 132 | 70 (57.4) | 81 (61.4) | LYMPH | 12 | 455 | 11 (91.7) | 110 (24.2) |
| LDL | 49 | 104 | 39 (79.6) | 58 (55.8) | WBC | 90 | 462 | 61 (67.8) | 158 (34.2) |
| TC | 75 | 145 | 66 (88.0) | 89 (61.4) |  |  |  |  |  |

**Note**: *G*_1_ and *G*_2_ are the number of genome-wide significance SNPs for the trait in the EAS or EUR population, respectively; *P*_1_ and *P*_2_ are the number of commonly associated SNPs (ccFDR<0.05) showing genome-wide significance (5×10^-8^) for the trait in the EAS or EUR population, respectively; and π_1_ and π_2_ are the corresponding proportion in each population.

#### Table S4. Results of MAGIC of effect size for all SNP in the EAS and EUR populations

| trait | *rm*_00_ | *fdr*_00_ | *rm*_10_ | *fdr*_10_ | *rm*_01_ | *fdr*_01_ | *rm*_11_ | *fdr*_11_ |
| --- | --- | --- | --- | --- | --- | --- | --- | --- |
| SCZ | 0.010 | 1.28×10^-8^ | -0.161 | 9.90×10^-1^ | -0.005 | 9.90×10^-1^ | -0.080 | 9.90×10^-1^ |
| RA | 0.016 | 1.47×10^-7^ | 0.339 | 2.98×10^-2^ | 0.290 | 3.74×10^-5^ | 0.462 | 4.78×10^-25^ |
| AF | 0.010 | 1.52×10^-2^ | -0.026 | 9.90×10^-1^ | 0.018 | 9.49×10^-1^ | 0.627 | 4.09×10^-21^ |
| T2D | 0.021 | 7.92×10^-34^ | 0.112 | 5.93×10^-1^ | 0.067 | 2.04×10^-1^ | 0.202 | 1.63×10^-2^ |
| COA | 0.012 | 9.65×10^-1^ | 0.507 | 2.02×10^-7^ | 0.047 | 5.89×10^-1^ | 0.427 | 1.53×10^-14^ |
| AOA | 0.015 | 9.65×10^-1^ | 0.294 | 5.10×10^-3^ | 0.129 | 9.74×10^-4^ | 0.525 | 1.18×10^-30^ |
| AD | 0.001 | 9.73×10^-1^ | 0.174 | 5.93×10^-1^ | -0.038 | 9.90×10^-1^ | 0.704 | 6.24×10^-16^ |
| BRC | 0.186 | 5.53×10^-6^ | 0.338 | 2.00×10^-16^ | 0.234 | 1.26×10^-6^ | 0.670 | 2.34×10^-19^ |
| IS | 0.008 | 9.73×10^-1^ | 0.517 | 1.22×10^-1^ | 0.591 | 1.06×10^-3^ | 0.364 | 1.35×10^-2^ |
| PCA | 0.007 | 2.67×10^-1^ | -0.033 | 9.90×10^-1^ | -0.304 | 9.90×10^-1^ | 0.680 | 9.71×10^-70^ |
| TL | 0.005 | 2.69×10^-1^ | -0.039 | 9.90×10^-1^ | 0.219 | 5.89×10^-1^ | 0.917 | 2.09×10^-137^ |
| BMI | 0.455 | 2.81×10^-94^ | 0.270 | 1.59×10^-3^ | 0.512 | 1.00×10^-124^ | 0.569 | 9.59×10^-41^ |
| height | 0.348 | 1.13×10^-34^ | 0.417 | 4.93×10^-18^ | 0.379 | 2.42×10^-39^ | 0.575 | 2.00×10^-126^ |
| DBP | 0.436 | 2.35×10^-45^ | 0.705 | 1.69×10^-5^ | 0.485 | 4.80×10^-82^ | 0.501 | 2.99×10^-30^ |
| SBP | 0.413 | 1.25×10^-55^ | 0.379 | 1.58×10^-2^ | 0.475 | 1.84×10^-85^ | 0.616 | 4.39×10^-79^ |
| PP | 0.472 | 1.22×10^-29^ | 0.511 | 1.59×10^-3^ | 0.452 | 2.47×10^-46^ | 0.763 | 1.01×10^-143^ |
| HDL | 0.598 | 5.91×10^-9^ | 0.422 | 1.59×10^-3^ | 0.740 | 1.61×10^-9^ | 0.670 | 4.77×10^-37^ |
| LDL | 0.528 | 3.71×10^-2^ | 0.086 | 7.97×10^-1^ | 0.122 | 5.88×10^-1^ | 0.484 | 1.44×10^-5^ |
| TC | 0.612 | 1.21×10^-2^ | 0.349 | 2.73×10^-2^ | 0.265 | 1.31×10^-2^ | 0.662 | 1.46×10^-9^ |
| TG | 0.615 | 2.83×10^-4^ | 0.416 | 4.16×50^-1^ | 0.325 | 1.45×10^-1^ | 0.768 | 1.35×10^-90^ |
| HbA1c | 0.014 | 1.20×10^-4^ | 0.274 | 1.10×10^-1^ | 0.501 | 3.27×10^-2^ | 0.628 | 8.63×10^-16^ |
| eGFR | 0.525 | 3.87×10^-28^ | 0.380 | 9.74×10^-10^ | 0.466 | 1.20×10^-40^ | 0.735 | 4.83×10^-109^ |
| AAM | 0.066 | 3.71×10^-2^ | -0.198 | 9.90×10^-1^ | -0.007 | 9.90×10^-1^ | -0.122 | 9.90×10^-1^ |
| ANM | 0.246 | 3.71×10^-2^ | -0.571 | 9.90×10^-1^ | 0.222 | 1.14×10^-1^ | 0.716 | 5.93×10^-15^ |
| PLT | 0.033 | 1.55×10^-26^ | 0.375 | 1.38×10^-6^ | 0.579 | 5.05×10^-69^ | 0.692 | 9.90×10^-190^ |
| RBC | 0.028 | 9.65×10^-1^ | 0.368 | 1.59×10^-3^ | 0.465 | 2.39×10^-36^ | 0.760 | 4.76×10^-195^ |
| MCV | 0.026 | 9.73×10^-1^ | 0.346 | 1.59×10^-3^ | 0.367 | 1.52×10^-24^ | 0.695 | 4.11×10^-193^ |
| HCT | 0.032 | 3.64×10^-1^ | 0.417 | 1.05×10^-2^ | 0.451 | 1.68×10^-33^ | 0.788 | 9.02×10^-98^ |
| MCH | 0.026 | 5.64×10^-1^ | 0.202 | 1.01×10^-1^ | 0.368 | 7.26×10^-24^ | 0.679 | 1.04×10^-146^ |
| MCHC | 0.018 | 7.13×10^-1^ | 0.218 | 9.94×10^-2^ | 0.478 | 6.86×10^-18^ | 0.744 | 3.26×10^-91^ |
| HGB | 0.033 | 9.65×10^-1^ | 0.847 | 1.67×10^-6^ | 0.429 | 1.05×10^-27^ | 0.786 | 6.12×10^-79^ |
| MONO | 0.019 | 9.73×10^-1^ | 0.330 | 1.69×10^-2^ | 0.455 | 2.09×10^-28^ | 0.727 | 1.06×10^-82^ |
| NEUT | 0.028 | 9.73×10^-1^ | 0.307 | 1.24×10^-1^ | 0.610 | 1.18×10^-50^ | 0.796 | 2.24×10^-132^ |
| EO | 0.016 | 9.73E-01 | 0.295 | 2.73×10^-2^ | 0.481 | 3.53×10^-29^ | 0.648 | 2.24×10^-42^ |
| BASO | 0.017 | 9.73E-01 | 0.398 | 1.22×10^-1^ | 0.297 | 2.14×10^-3^ | 0.698 | 1.48×10^-46^ |
| LYMPH | 0.027 | 9.73E-01 | 0.987 | 1.08×10^-10^ | 0.613 | 9.06×10^-73^ | 0.662 | 1.02×10^-23^ |
| WBC | 0.033 | 9.73E-01 | 0.243 | 2.72×10^-2^ | 0.582 | 2.53×10^-36^ | 0.813 | 2.00×10^-16^ |

**Note**: "no" column means the number of the trait; *rm*_00_ is the marginal genetic correlation of null SNPs effect sizes; *rm*_10_ is the marginal genetic correlation of EAS-specific SNPs effect sizes; *rm*_01_ is the marginal genetic correlation of EUR-specific SNPs effect sizes; *rm*_11_ is the marginal genetic correlation of population-common SNPs effect sizes; *fdr*_00_ is the FDR q-value of null SNPs effect sizes; *fdr*_10_ is the FDR q-value of EAS-specific SNPs effect sizes; *fdr*_01_ is the FDR q-value of EUR-specific SNPs effect sizes; *fdr*_11_ is the FDR q-value of population-common SNPs effect sizes.

#### Table S5. Results of Cochran's Q test of effect size for shared associated SNP (ccFDR<0.05) in the EAS and EUR populations

| trait | *f*_11_ | Q (%) | trait | *f*_11_ | Q (%) |
| --- | --- | --- | --- | --- | --- |
| SCZ | 605 | 82 (13.6) | TG | 181 | 42 (23.2) |
| RA | 881 | 342 (38.8) | HbA1c | 106 | 40 (37.7) |
| AF | 240 | 43 (17.9) | eGFR | 933 | 199 (21.3) |
| T2D | 1904 | 232 (12.2) | AAM | 340 | 180 (52.9) |
| COA | 421 | 113 (26.8) | ANM | 85 | 17 (20.0) |
| AOA | 432 | 126 (29.2) | PLT | 1,205 | 174 (14.4) |
| AD | 80 | 28 (35.0) | RBC | 890 | 83 (9.3) |
| BRC | 71 | 12 (16.9) | MCV | 1273 | 204 (16.0) |
| IS | 39 | 12 (30.8) | HCT | 457 | 51 (11.2) |
| PCA | 293 | 63 (21.5) | MCH | 1,184 | 200 (16.9) |
| TL | 42 | 3 (7.1) | MCHC | 338 | 64 (18.9) |
| BMI | 767 | 118 (15.4) | HGB | 339 | 44 (13.0) |
| height | 2616 | 384 (14.7) | MONO | 440 | 86 (19.5) |
| DBP | 468 | 102 (21.8) | NEUT | 309 | 42 (13.6) |
| SBP | 642 | 142 (22.1) | EO | 334 | 55 (16.5) |
| PP | 409 | 52 (12.7) | BASO | 200 | 46 (23.0) |
| HDL | 226 | 59 (26.1) | LYMPH | 263 | 41 (15.6) |
| LDL | 177 | 48 (27.1) | WBC | 555 | 68 (12.3) |
| TC | 275 | 67 (24.4) |  |  |  |

**Note**: *f*_11_ is the number of shared associated SNPs; Q (%) is the number (or proportion) of identified SNPs with heterogeneous effect.

#### Table S6. Summary information for traits analyzed in the present study

| trait | *k* | East Asian (EAS) | | |  | European (EUR) | | |
| --- | --- | --- | --- | --- | --- | --- | --- | --- |
|  |  | *n*_1_ | *k*_1_ | ref |  | *n*_1_ | *k*_2_ | ref |
| binary trait | | | | | | | | |
| SCZ (schizophrenia) | 458,236 | 55,416 | 279,265  107,727 | [1] |  | 79,925 | 251,421 | [2] |
| RA (rheumatoid arthritis) | 203,961 | 15,273 | 107,727 | [3] |  | 43,290 | 111,682 | [3] |
| AF (atrial fibrillation) | 236,293  475,656 | 25,445 | 115,869  284,894 | [4] |  | 228,221 | 139,158 | [5] |
| T2D (type 2 diabetes) | 475,656  250,069 | 254,373 | 284,894 | [6] |  | 272,026 | 272,261 | [7] |
| COA (childhood onset asthma) | 250,069 | 31,577 | 133,291 | [8] |  | 53,370 | 138,912 | [9] |
| AOA (adult onset asthma） | 249,871 | 31,577 | 133,256  138,250 | [8] |  | 97,691 | 138,801 | [9] |
| AD (atopic dermatitis) | 274,668 | 9,433 | 138,250  201,936 | [8] |  | 61,737 | 161,490 | [10] |
| BRC (breast cancer) | 337,234 | 12,788 | 201,936 | [11] |  | 227,688 | 190,473 | [12] |
| IS (ischemic stroke) | 246,666  247,201 | 64,738 | 130,088 | [8] |  | 147,590 | 137,946 | [13] |
| PCA (prostate cancer) | 247,201 | 20,562 | 129,257  129,257 | [8] |  | 137,462 | 140,161 | [14] |
| continuous trait |  |  |  |  |  |  |  |  |
| TL (telomere length) | 125,043 | 23,096 | 66,344 | [15] |  | 37,684 | 69,821 | [16] |
| BMI (body mass index) | 118,899 | 158,284 | 64,877 | [17] |  | 681,275 | 65,304 | [18] |
| Height | 139,126 | 159,095 | 77,847 | [19] |  | 693,529 | 76,625 | [18] |
| DBP (diastolic blood pressure) | 214,705 | 136,615 | 112,966 | [19] |  | 757,601 | 119,032 | [20] |
| SBP (systolic blood pressure) | 213,840 | 136,597 | 112,675 | [19] |  | 757,601 | 118,211 | [20] |
| PP (pulse pressure) | 213,862 | 136,249 | 112,625 | [19] |  | 757,601 | 118,206 | [20] |
| HDL (high density lipoprotein cholesterol) | 122,159 | 70,657 | 66,853 | [19] |  | 95,123 | 66,996 | [21] |
| LDL (low density lipoprotein cholesterol) | 121,447 | 72,866 | 66,541 | [19] |  | 90,422 | 66,460 | [21] |
| TC (total cholesterol) | 122,106 | 128,305 | 66,877 | [19] |  | 95,358 | 66,958 | [21] |
| TG (triglyceride) | 121,414 | 105,597 | 66,485 | [19] |  | 91,598 | 66,423 | [21] |
| HbA1c (hemoglobin) | 124,962 | 42,790 | 68,163 | [19] |  | 123,665 | 68,875 | [22] |
| eGFR (estimated glomerular filtration rate) | 236,693 | 143,658 | 120,334 | [19] |  | 765,348 | 135,987 | [23] |
| AAM (age at menarche) | 319,439 | 67,029 | 185,610 | [19] |  | 329,345 | 172,009 | [24] |
| ANM (age at menopause) menopause) | 137,069 | 43,861 | 79,292 | [19] |  | 69,360 | 91,113 | [25] |
| PLT (platelet count) | 222,492 | 108,208 | 115,817 | [19] |  | 173,480 | 125,360 | [26] |
| RBC (red blood cell count; ) | 222,589 | 108,794 | 115,897 | [19] |  | 173,480 | 125,446 | [26] |
| MCV (mean corpuscular volume) | 222,587 | 108,256 | 115,813 | [19] |  | 173,480 | 125,511 | [26] |
| HCT (hematocrit) | 222,528 | 108,757 | 115,891 | [19] |  | 173,480 | 125,388 | [26] |
| MCH (mean corpuscular hemoglobin) | 222,441 | 108,054 | 115,785 | [19] |  | 173,480 | 125,343 | [26] |
| MCHC (mean corpuscular hemoglobin concentration) | 222,580 | 108,728 | 115,904 | [19] |  | 173,480 | 125,451 | [26] |
| HGB (hemoglobin concentration) | 222,563 | 108,769 | 115,915 | [19] |  | 173,480 | 125,382 | [26] |
| MONO (monocyte count) | 222,622 | 62,076 | 115,813 | [19] |  | 173,480 | 125,532 | [26] |
| NEUT (neutrophil count) | 222,435 | 62,076 | 115,820 | [19] |  | 173,480 | 125,319 | [26] |
| EO (eosinophil count) | 222,395 | 62,076 | 115,811 | [19] |  | 173,480 | 125,368 | [26] |
| BASO (basophil count) | 222,525 | 62,076 | 115,917 | [19] |  | 173,480 | 125,330 | [26] |
| LYMPH (lymphocyte count) | 222,490 | 62,076 | 115,920 | [19] |  | 173,480 | 125,287 | [26] |
| WBC (white blood cell count) | 222,550  222,550 | 107,964 | 115,908 | [19] |  | 173,480 | 125,382 | [26] |

**Note**: *k* is the number of combined SNPs after pruning, *k*_1_ and *k*_2_ are the number of independent SNPs in the each population, *n*_1_ and *n*_2_ are the sample size of the EAS or EUR population, which is the original sample size for continuous traits and is the effective sample size for binary traits computed via 4/(1/*n*_case_+1/*n*_control_) [27].

#### Table S7. Number of case and control for binary traits in the EAS and EUR populations

| trait | EAS | |  | EUR | |
| --- | --- | --- | --- | --- | --- |
|  | *n (case/control)* | ref |  | *n (case/control)* | ref |
| SCZ | 55,416 (22,778/35,362) | [1] |  | 79,925 (36,989/43,456) | [2] |
| RA | 15,273 (4,873/17,642) | [3] |  | 43,290 (14,361/43,923) | [3] |
| AF | 25,445 (8,180/28,612) | [4] |  | 228,221 (60,620/970,216) | [5] |
| T2D | 254,373 (77,41/356,122) | [6] |  | 272,026 (74,124/824,006) | [7] |
| COA | 31,577 (8,216/201,592) | [8] |  | 53,370 (13,962/300,671) | [9] |
| AOA | 31,577 (8,216/201,592) | [8] |  | 97,691 (26,582/300,671) | [9] |
| AD | 9,433 (2,385/209,651) | [8] |  | 61,737 (18,900/84,166) | [10] |
| BRC | 12,788 (6,269/6,524) | [11] |  | 227,688 (122,977/105,974) | [12] |
| IS | 64,738 (17,671/192,383) | [8] |  | 147,590 (40,585/406,111) | [13] |
| PCA | 20,562 (5,408/103,939) | [8] |  | 137,462 (60,165/80,141) | [28] |

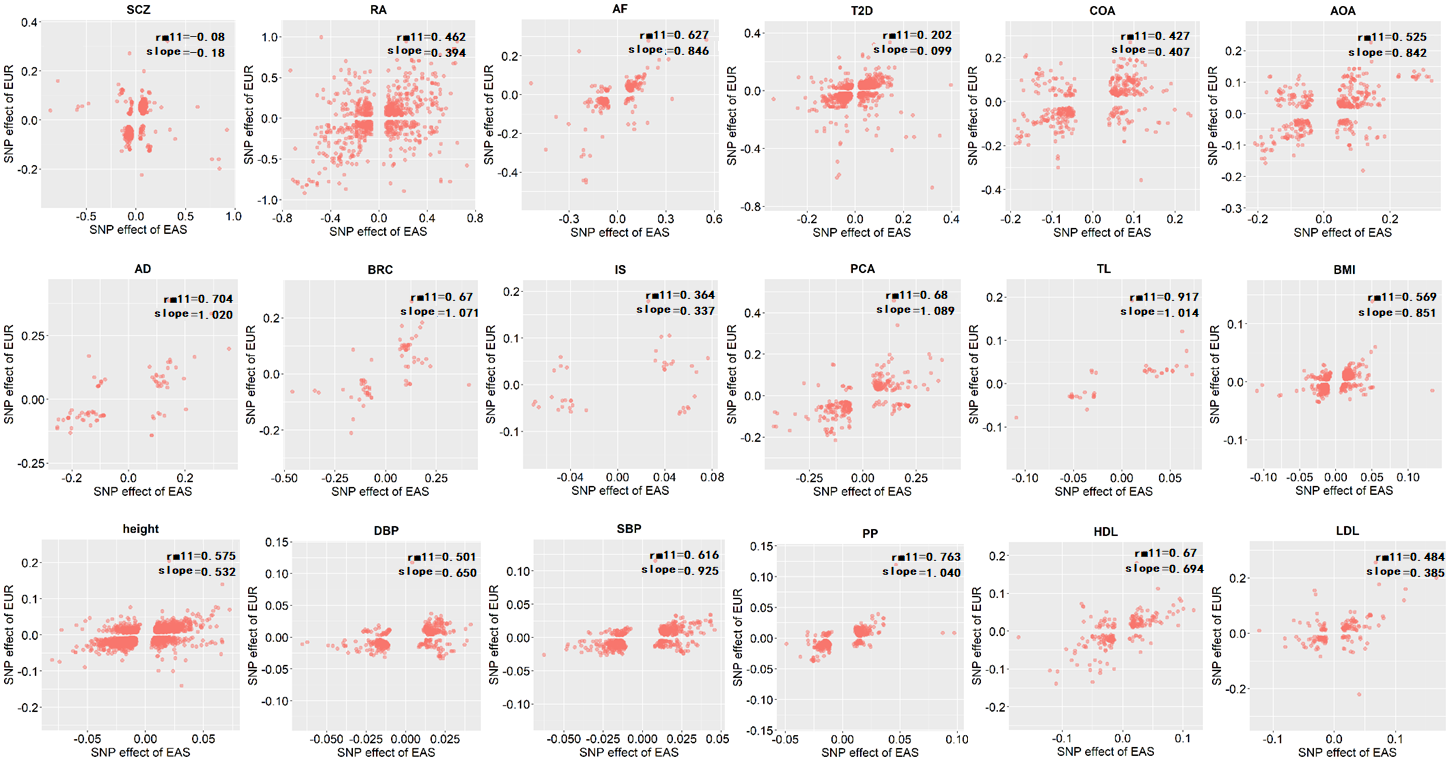

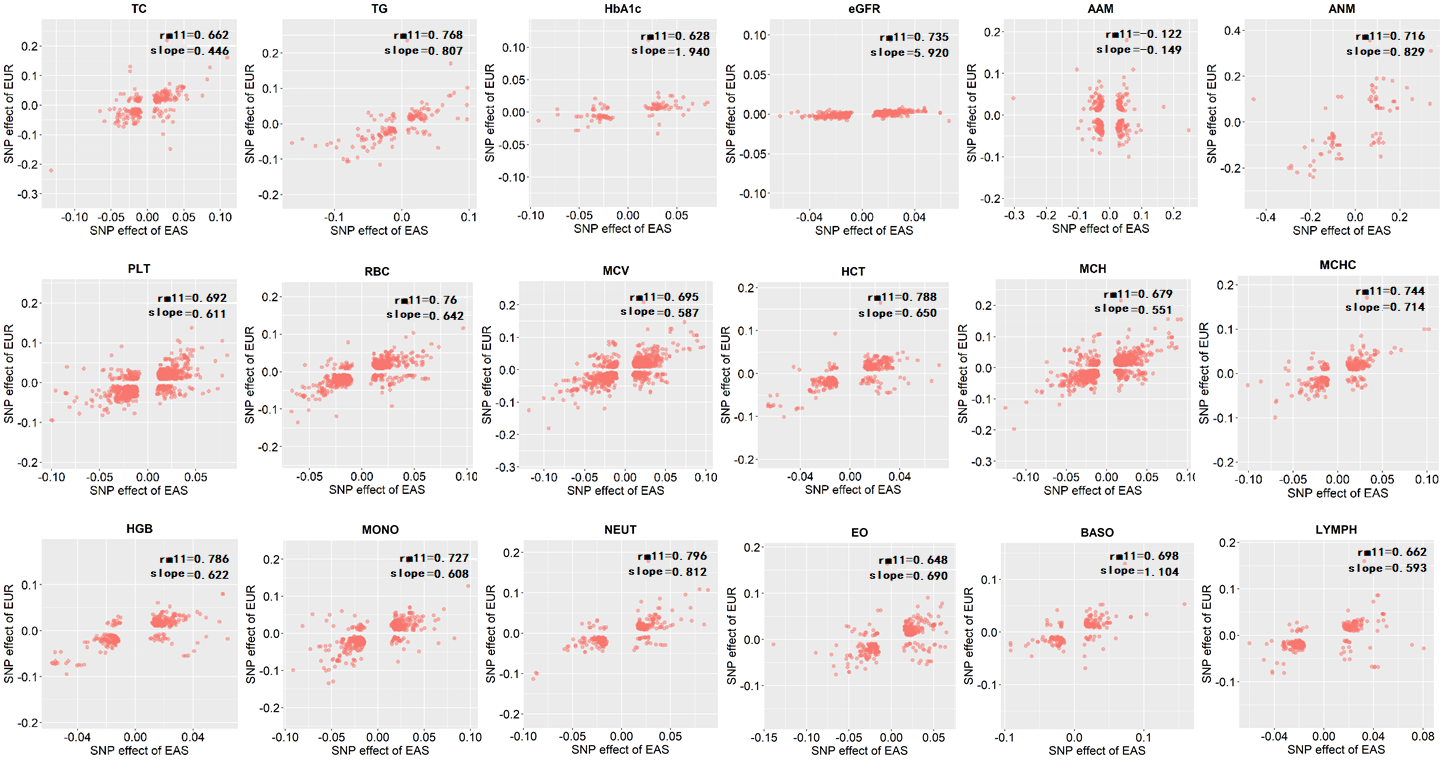

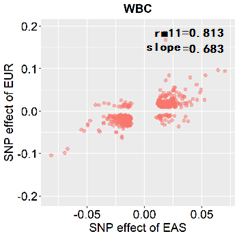

#### **Figure S1**. Relationship for effect size of shared associated SNPs for traits in the EAS and EUR populations. Here *rm*_11_ is the marginal genetic correlation of population-common SNPs effect sizes; slope is the regression coefficient by regressing population-common SNP effect sizes in one population on those in another population.

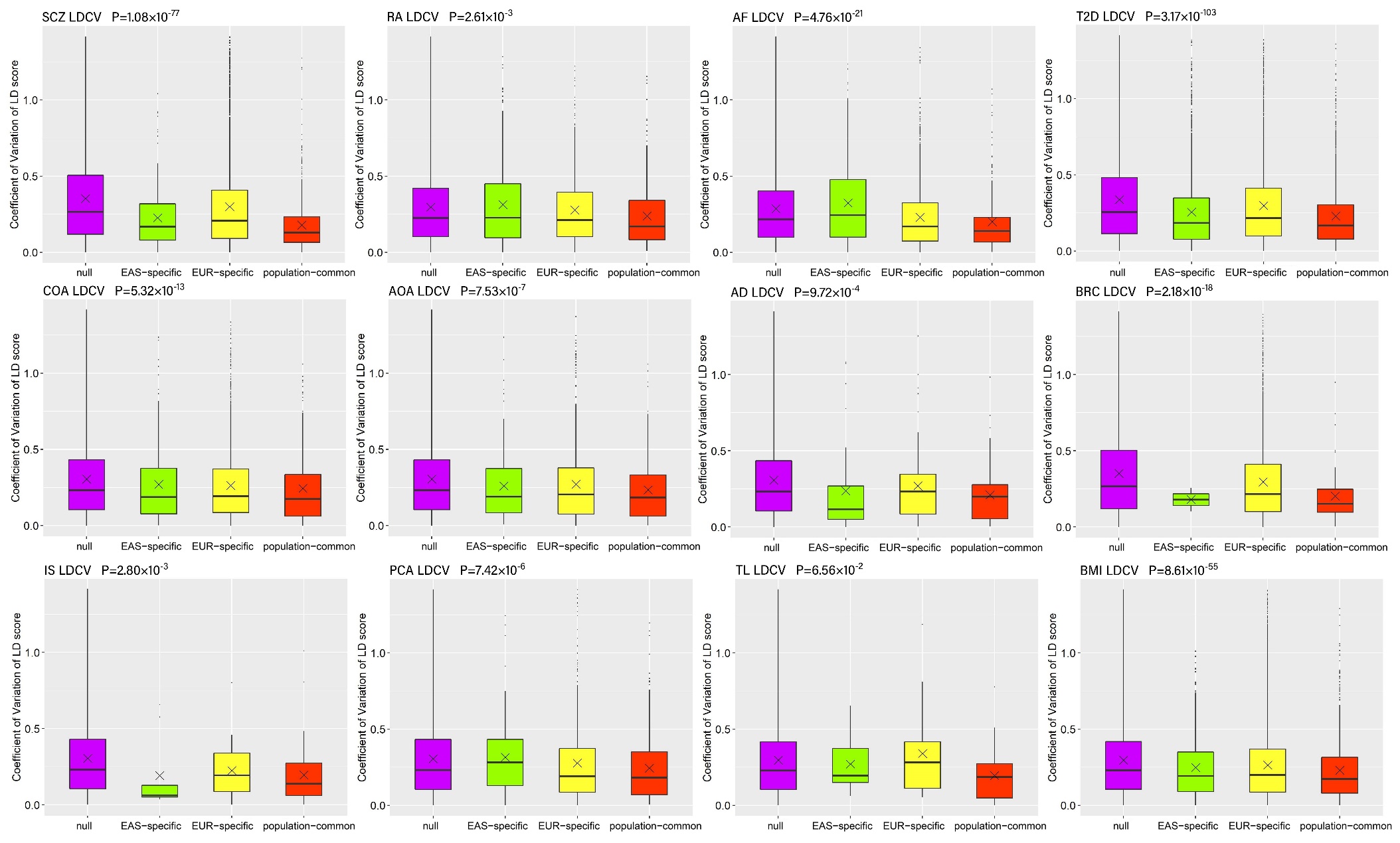

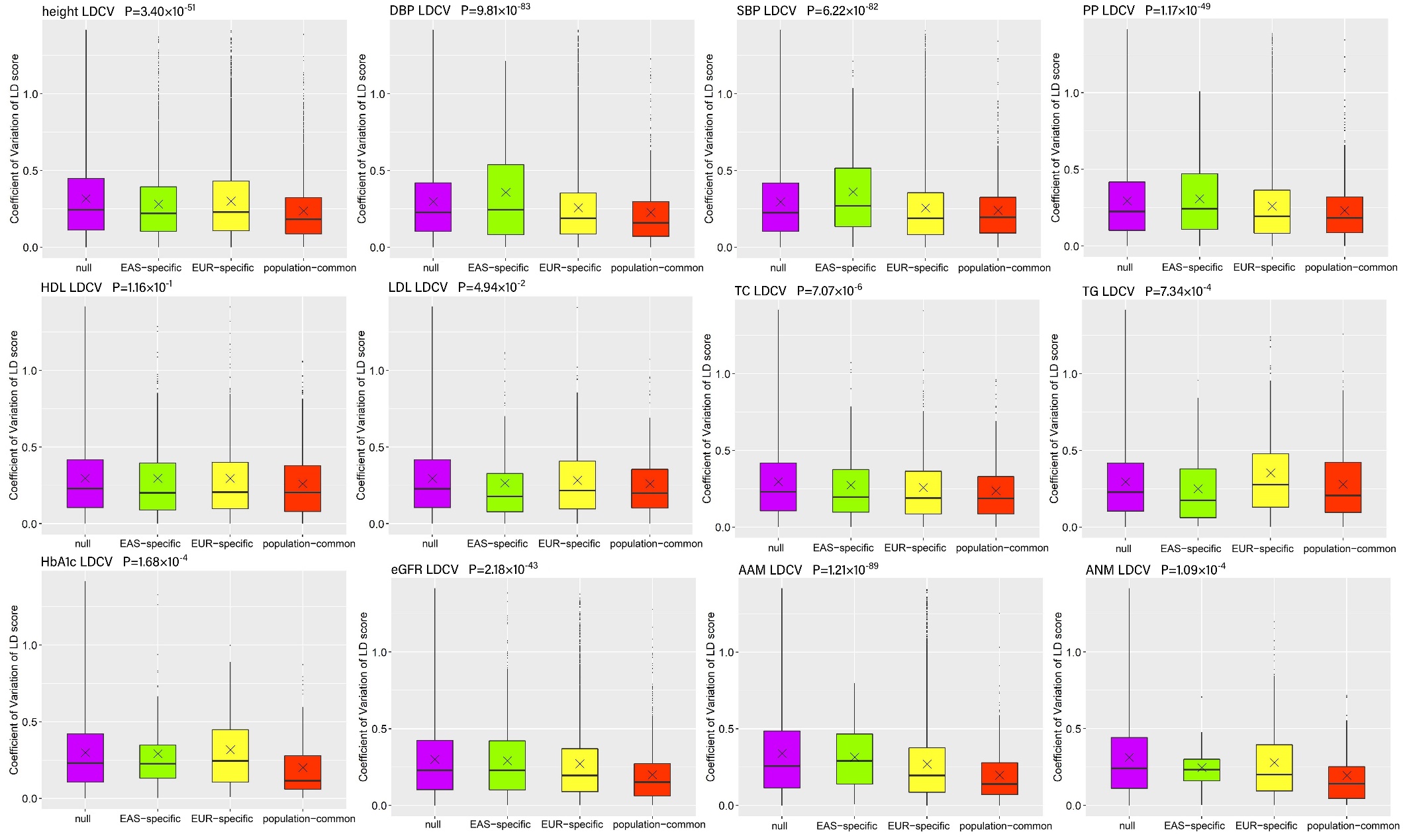

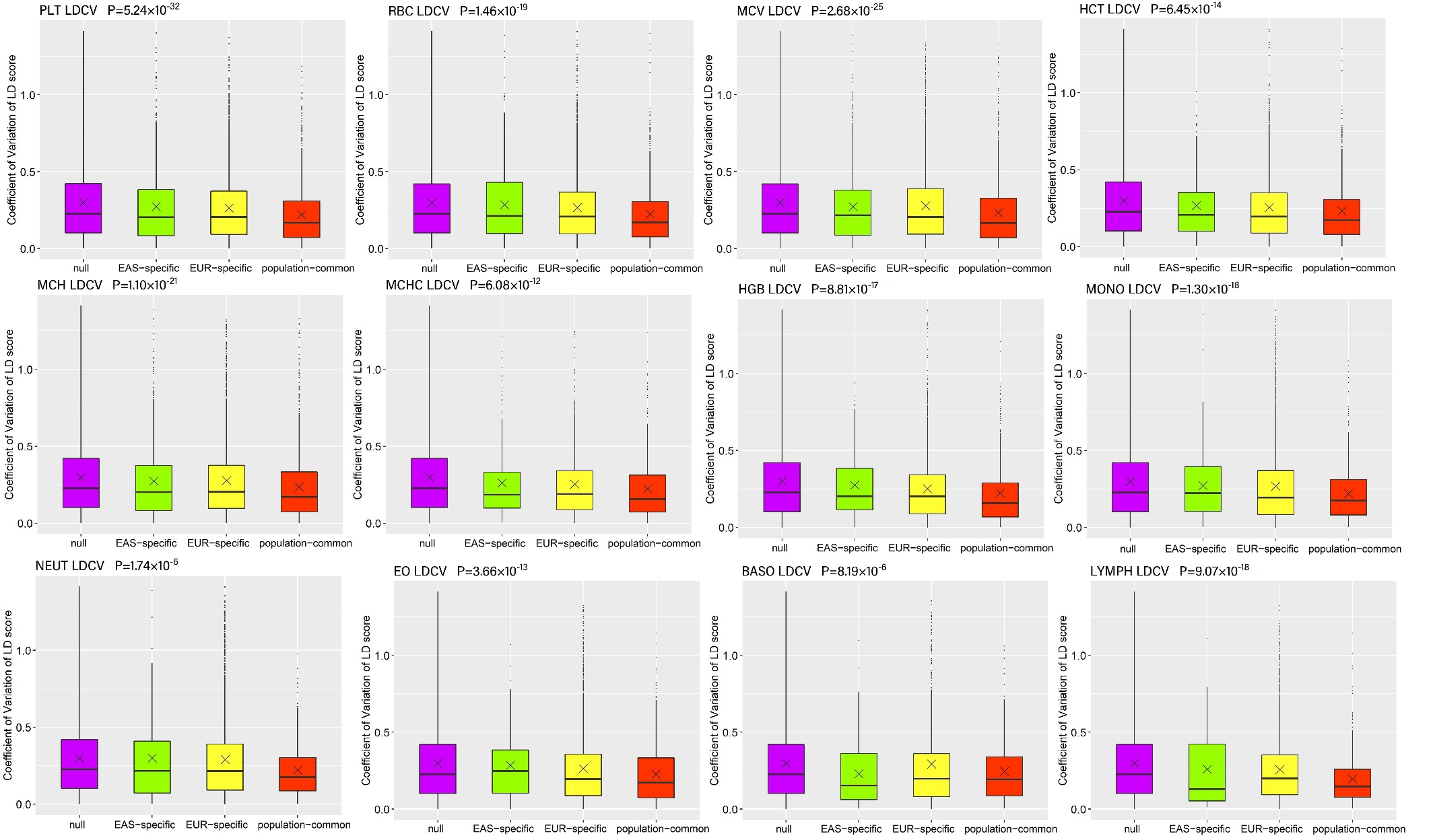

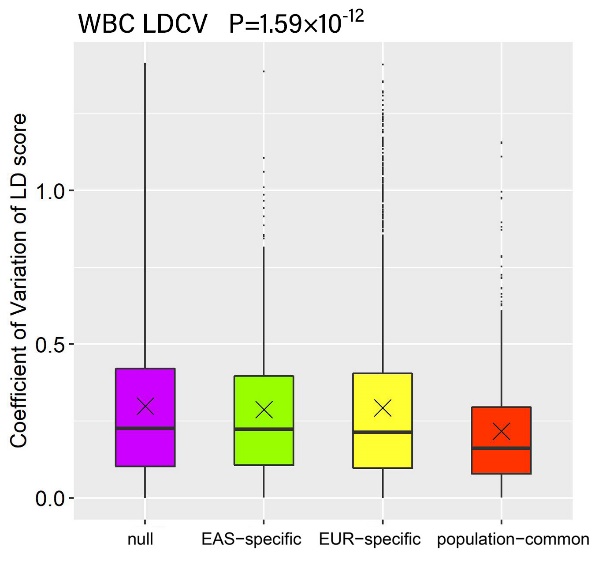

#### **Figure S2.** Distribution of LDCV for SNPs in the four diverse groups for each analyzed trait. The shown *P* value in each panel is available in terms of the Kruskal test for LDCV among the four groups. × means the median.

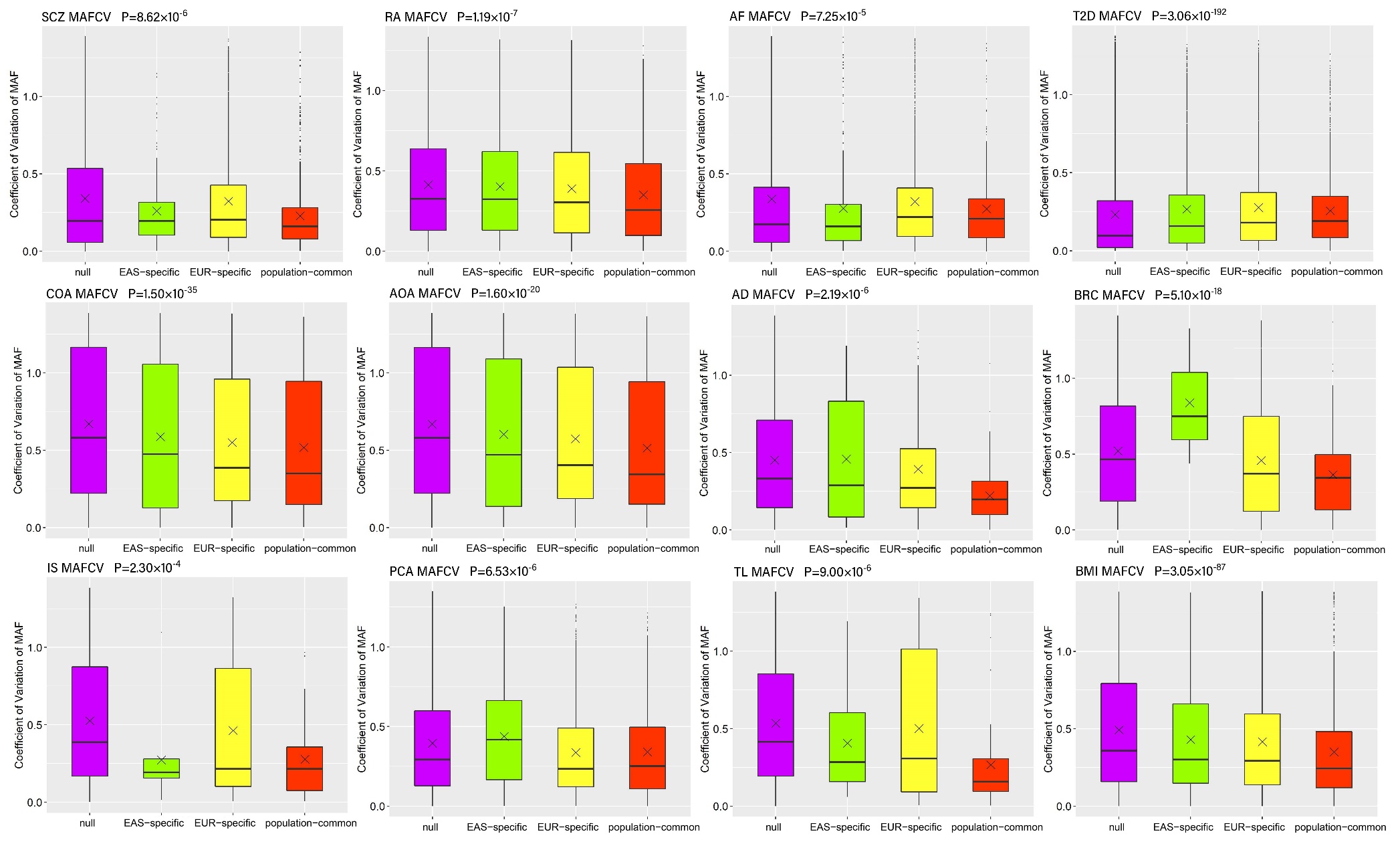

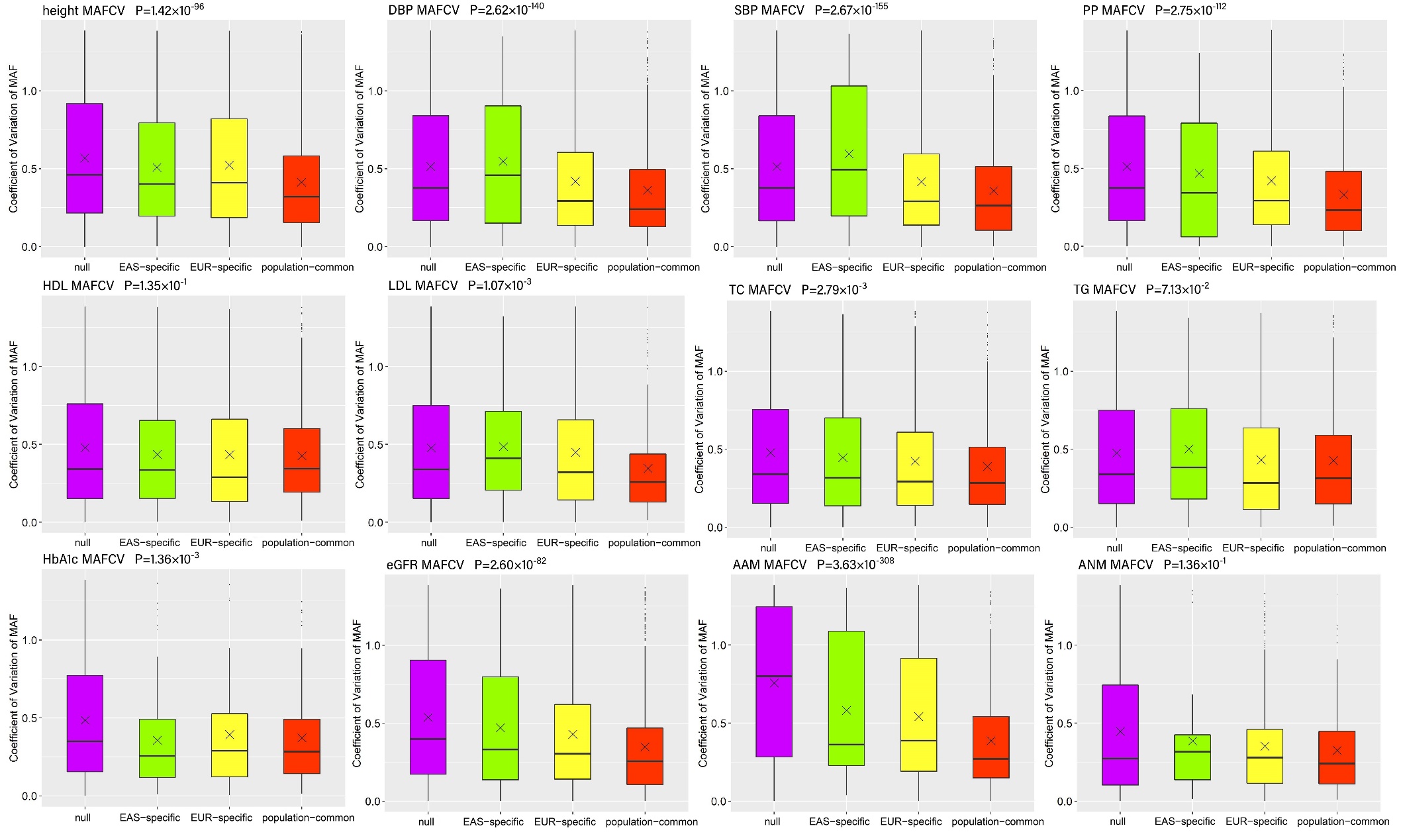

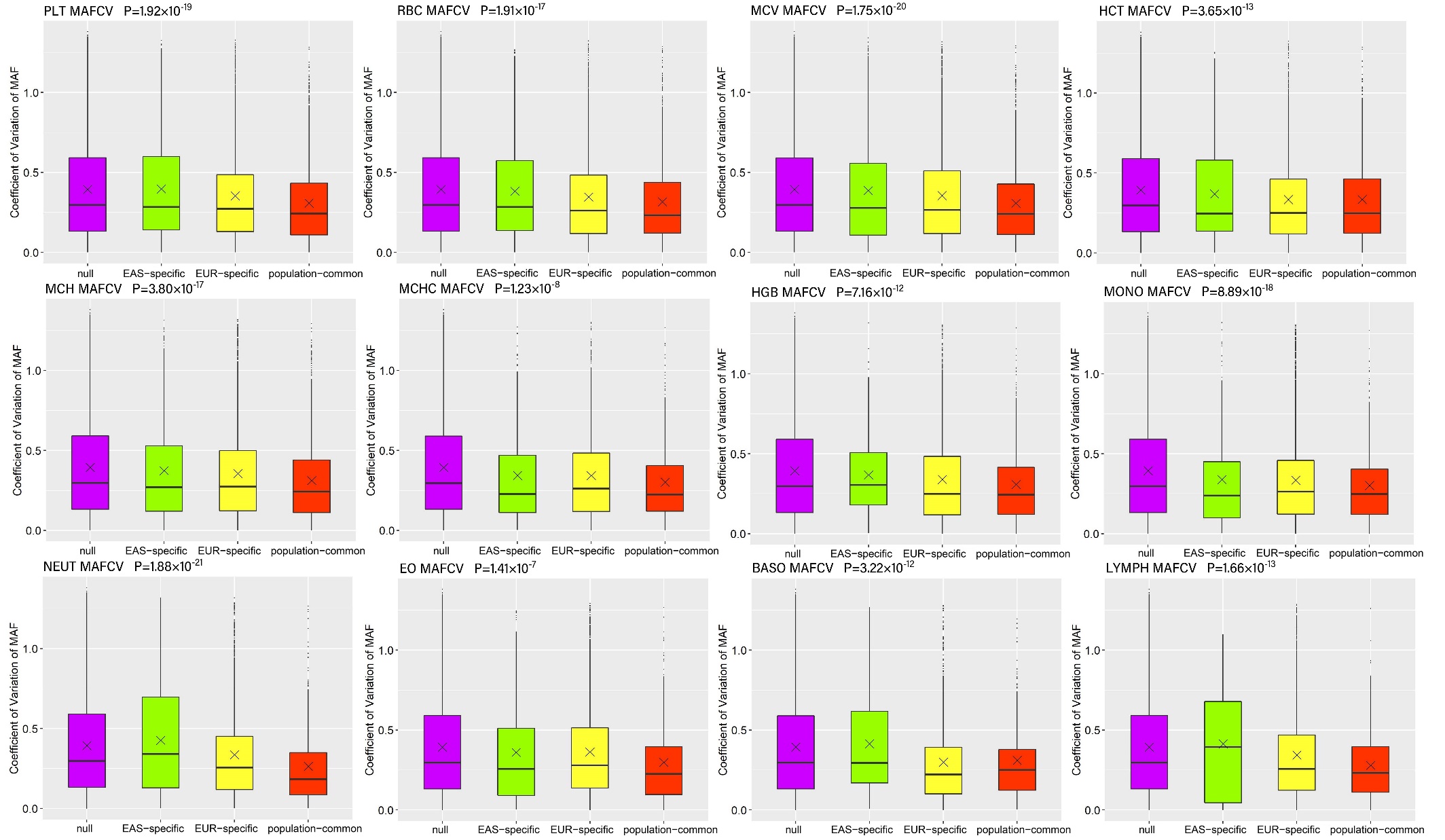

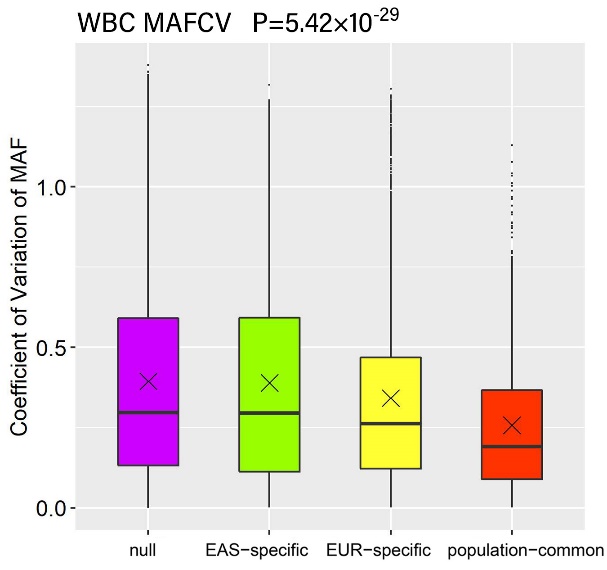

#### **Figure S3.** Distribution of MAFCV for SNPs in the four diverse groups for each analyzed trait. The shown *P* value in each panel is available in terms of the Kruskal test for MAFCV among the four groups. × means the median.

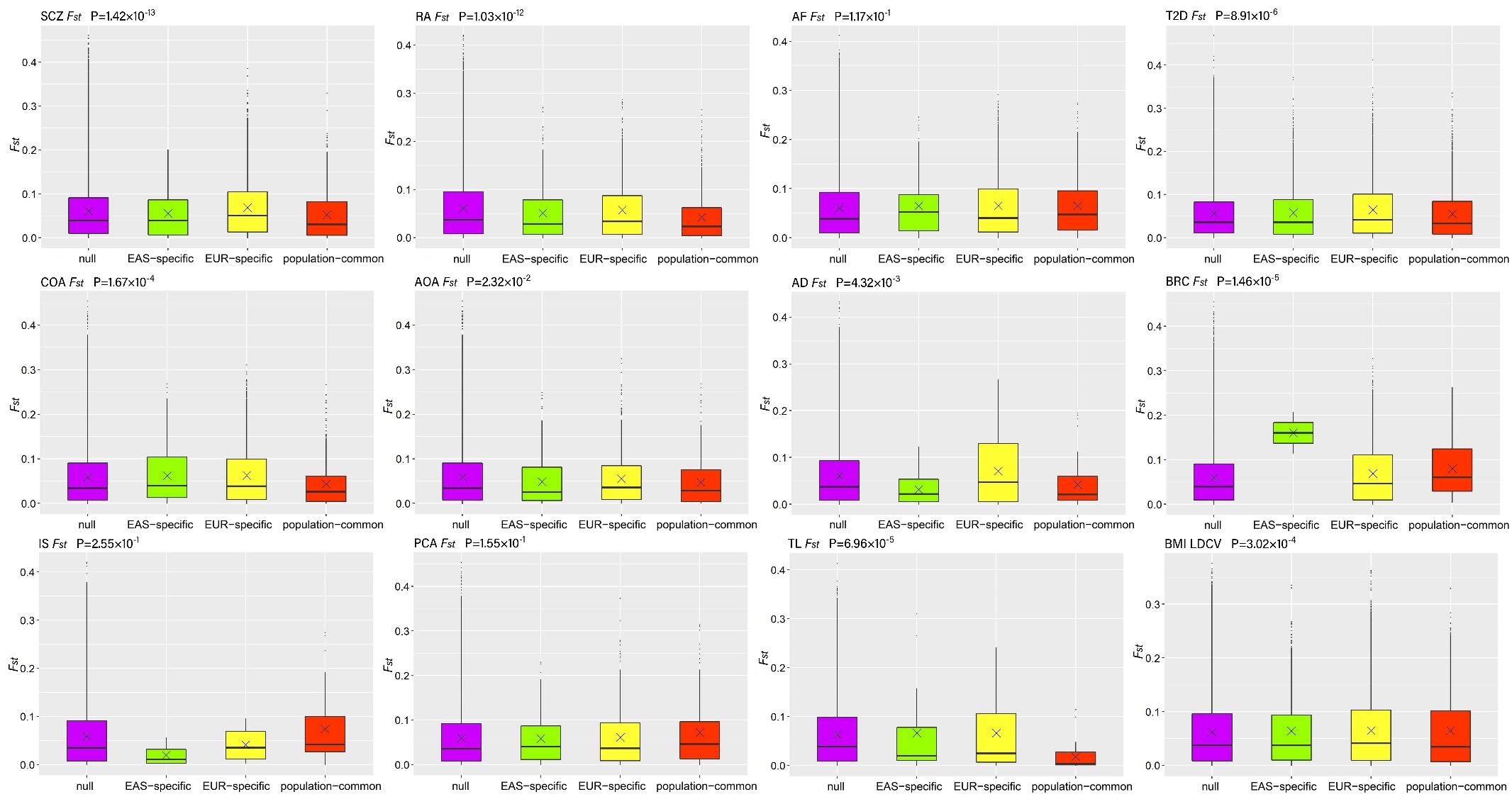

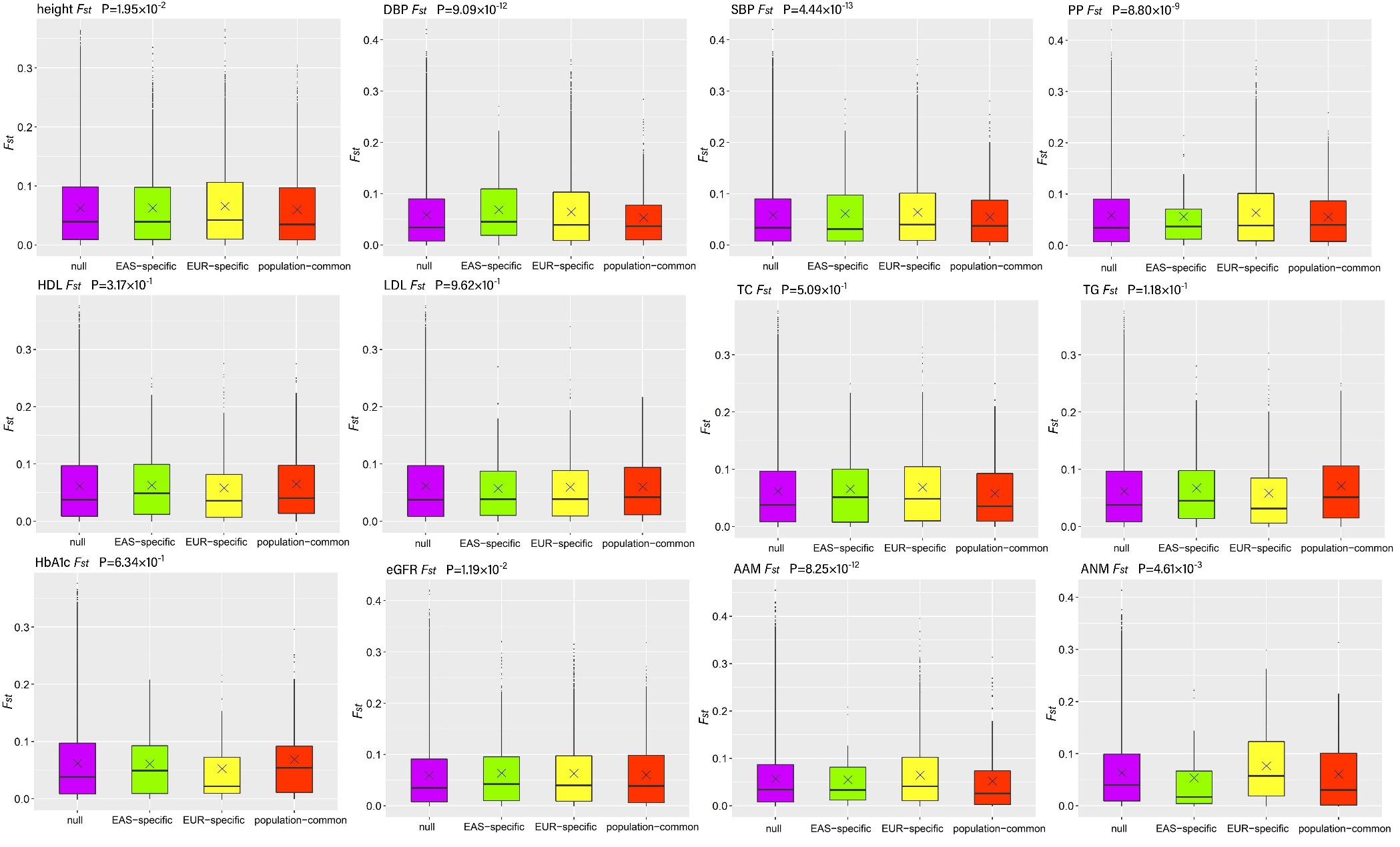

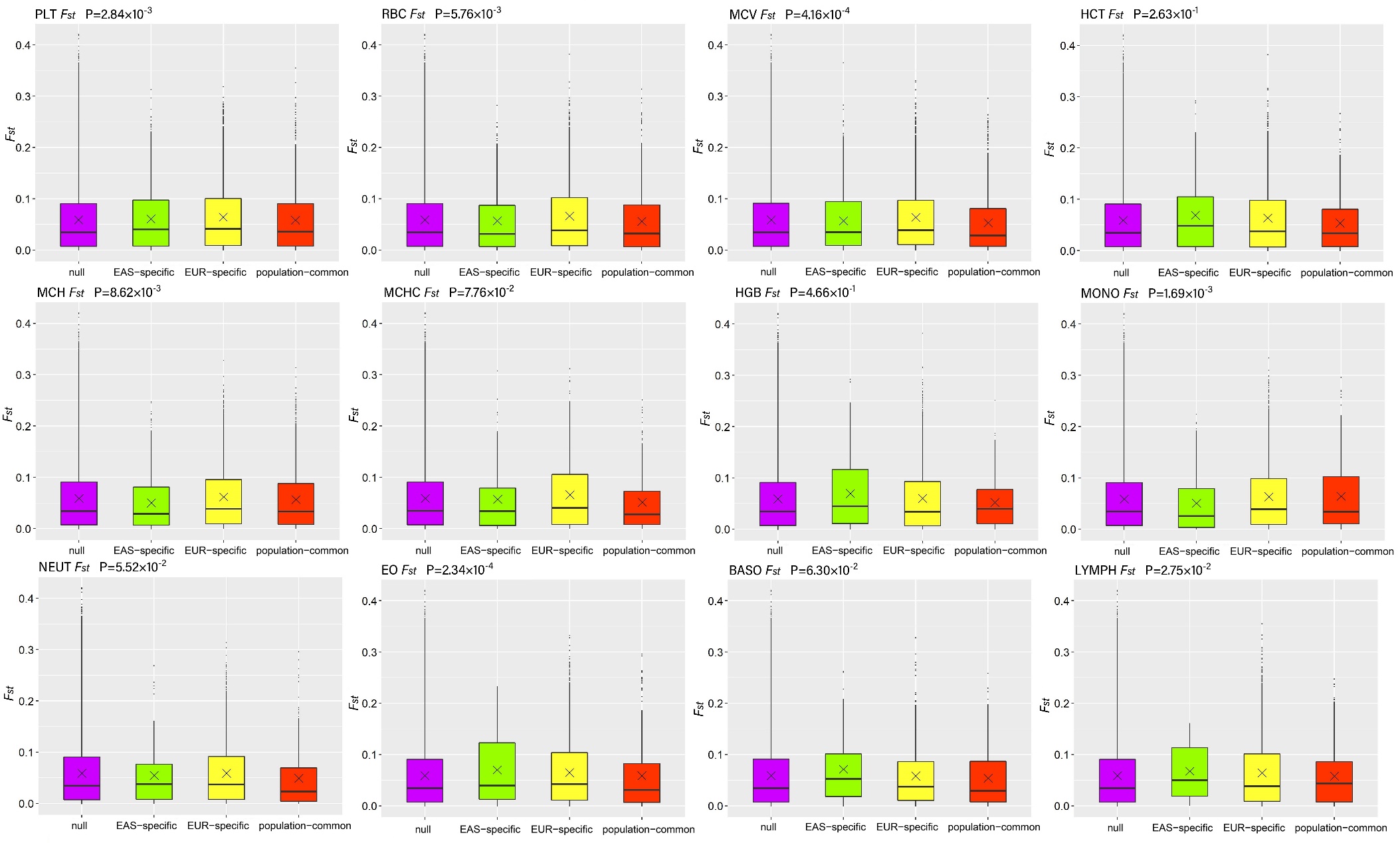

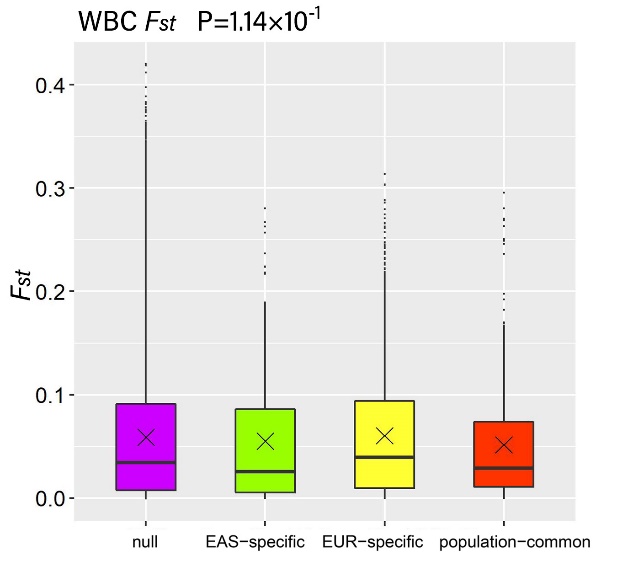

#### **Figure S4.** Distribution of *F_st_* for SNPs in the four diverse groups for each analyzed trait. The shown *P* value in each panel is available in terms of the Kruskal test for *F_st_* among the four groups. × means the median.

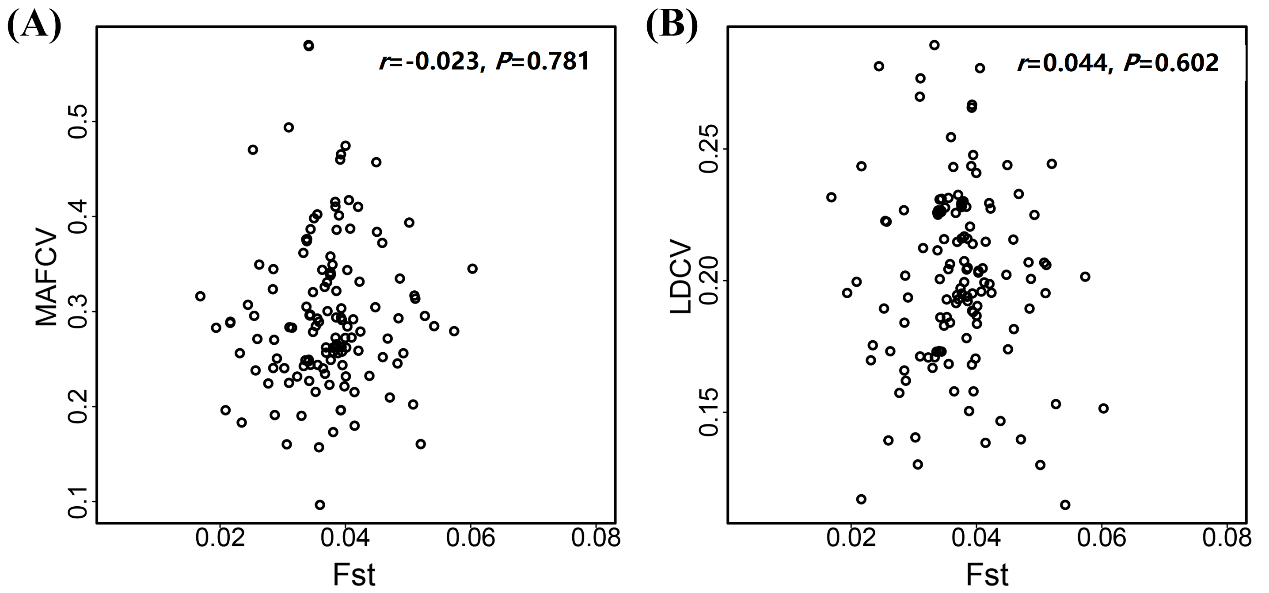

#### **Figure S5.** (**A**) Relationship between average *F_st_* and average MAFCV across all analyzed traits. (**B**) Relationship between average *F_st_* and average LDCV across all analyzed traits.

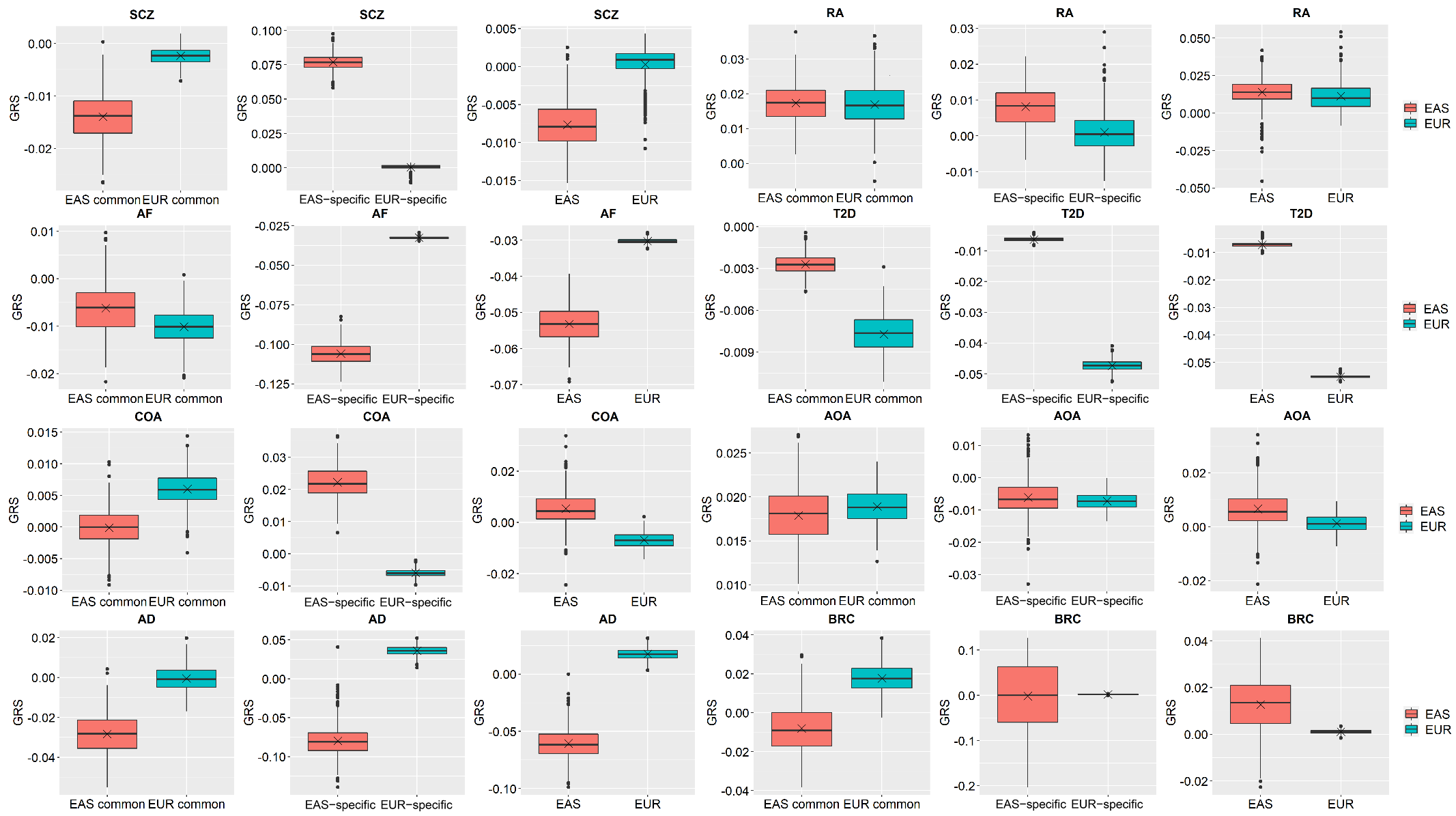

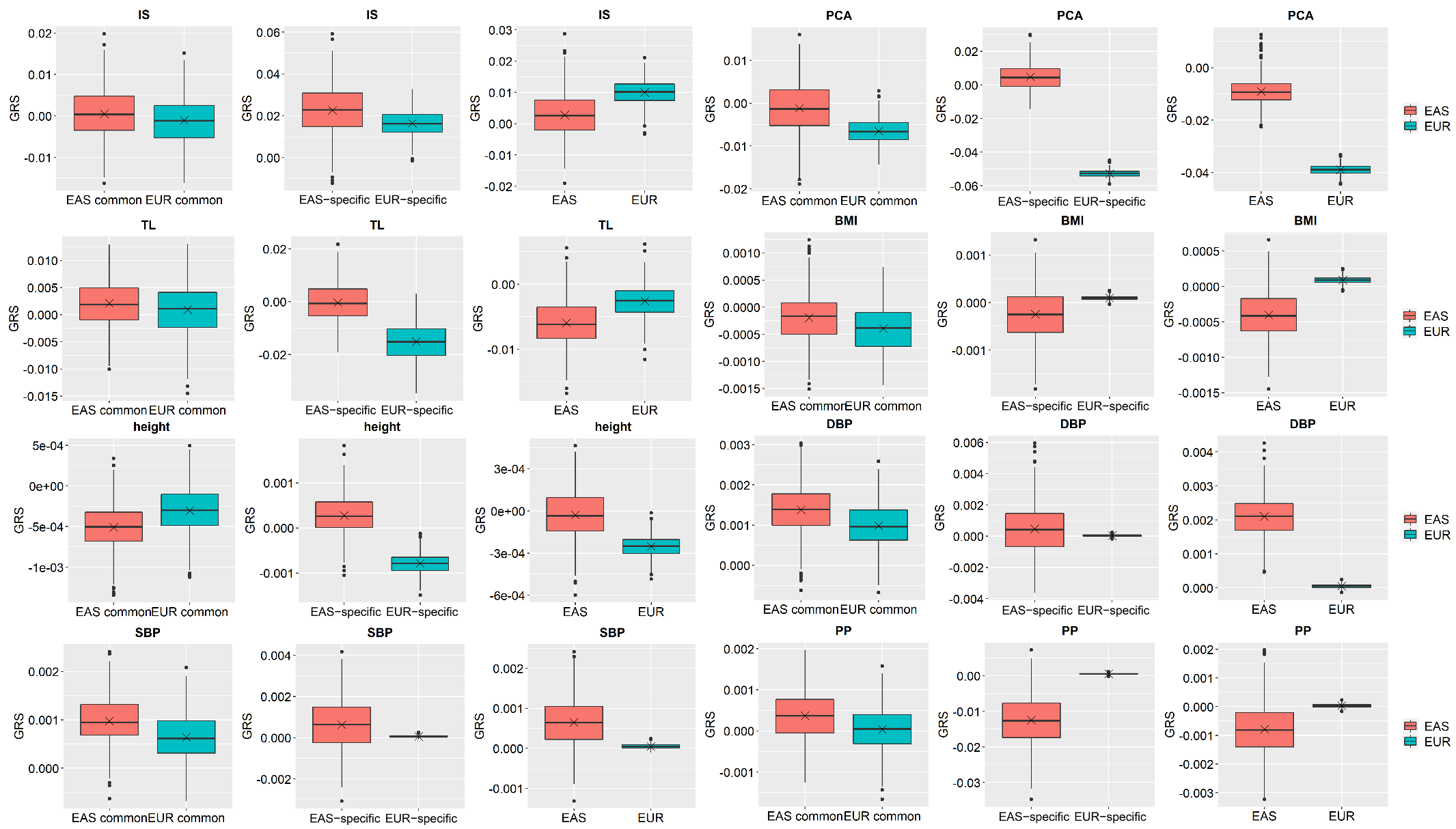

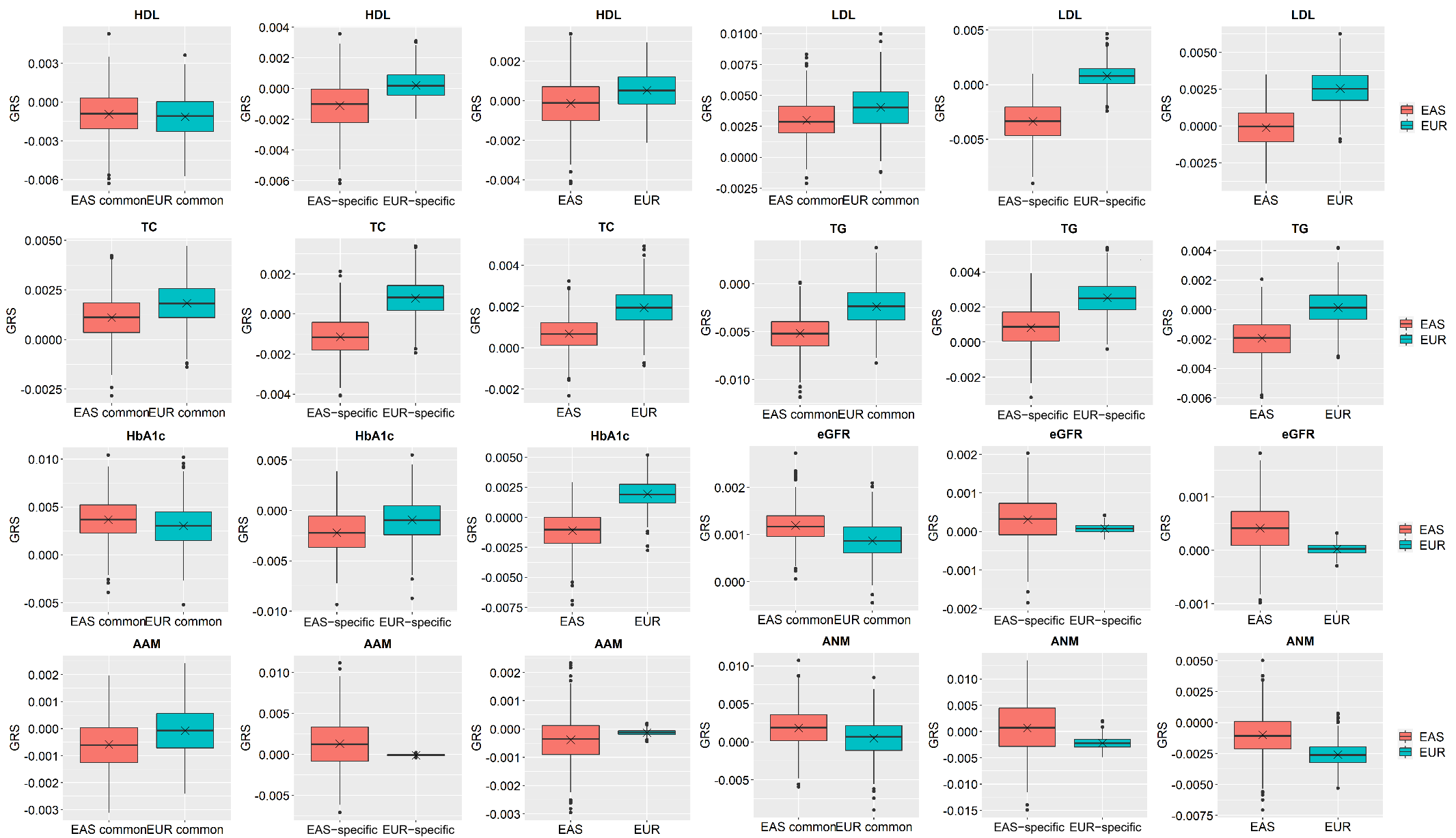

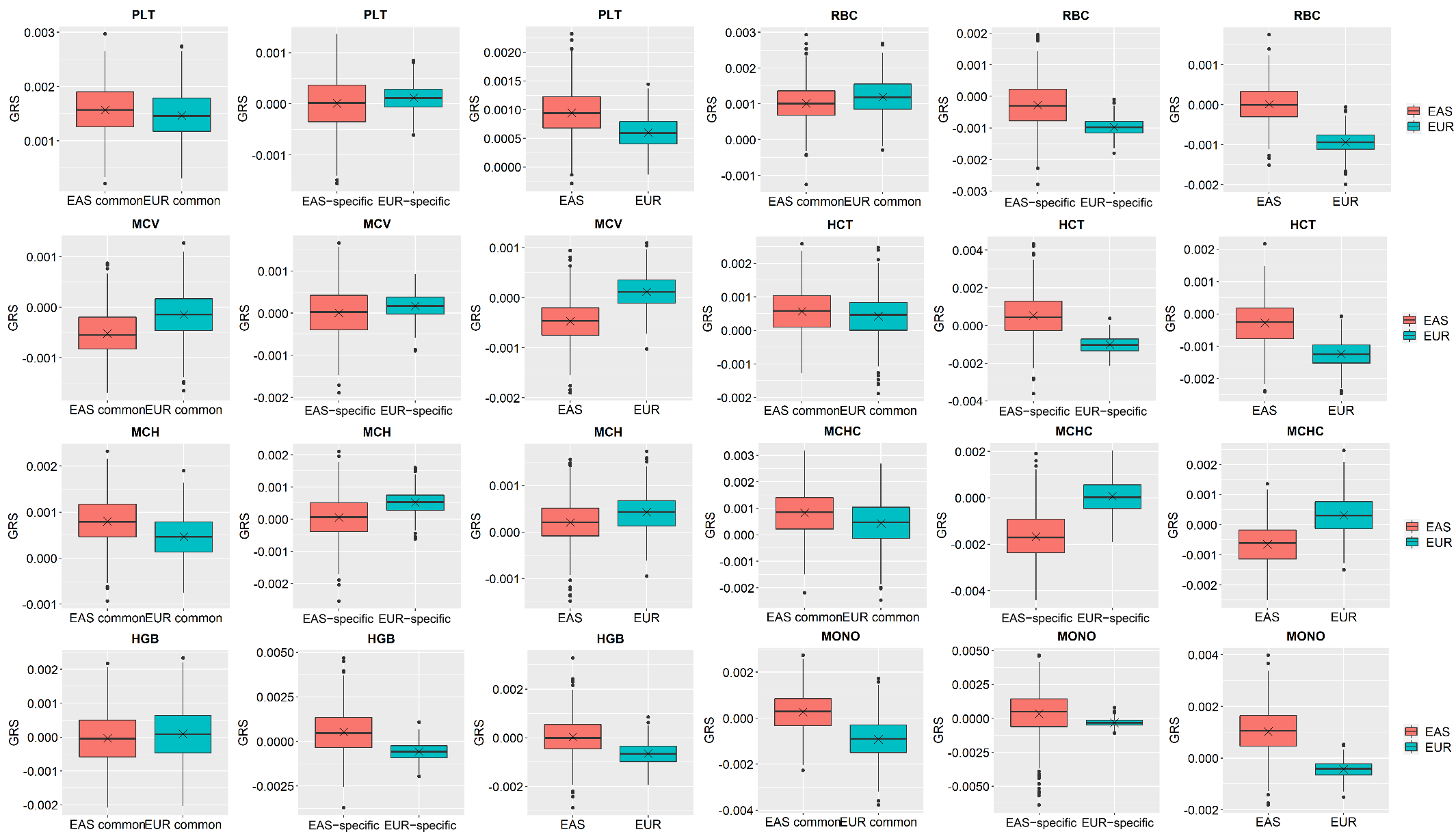

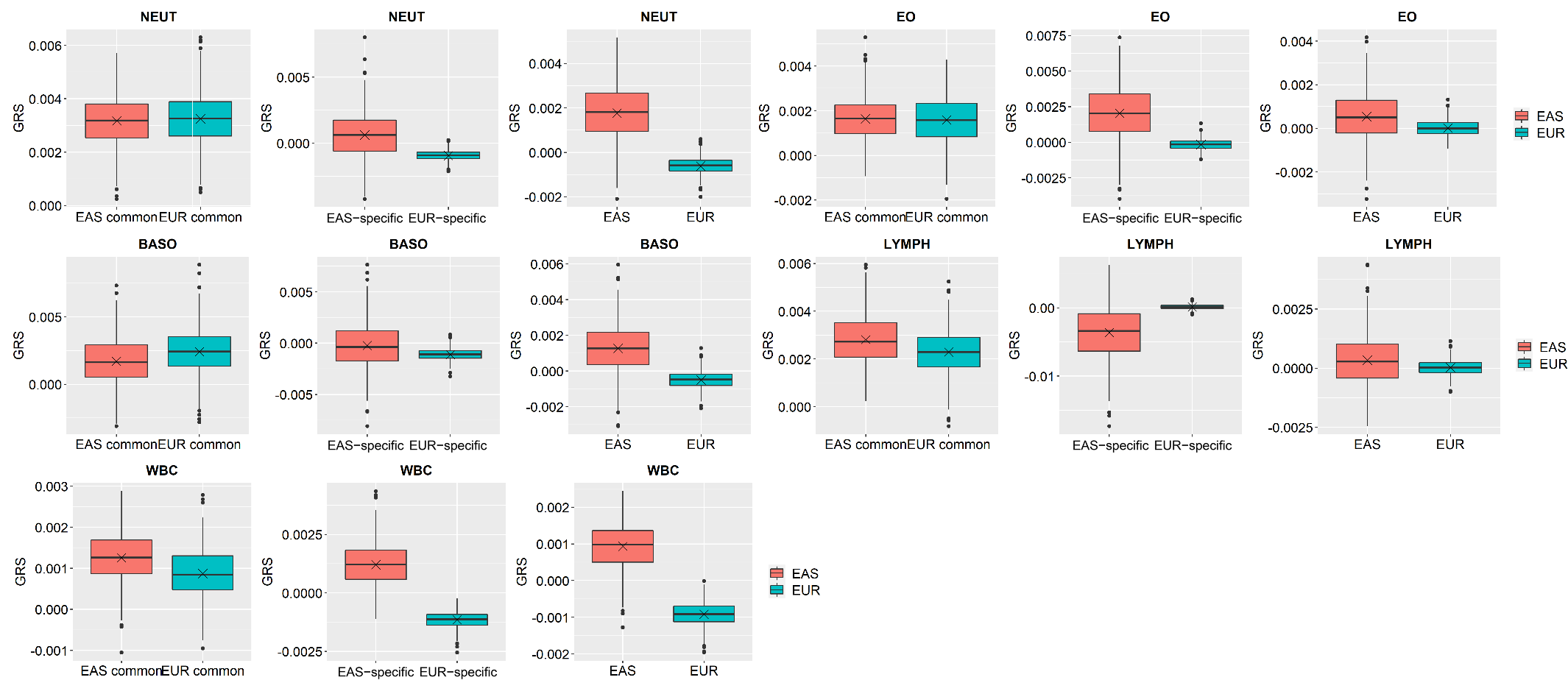

#### **Figure S6**. Distribution of GRS for each analyzed trait calculated with population-common SNPs (the first one), population-specific SNPs (the second one) or all trait-associated SNPs (the third one). The average of the genetic risk score for each disease across individuals is shown in each panel.

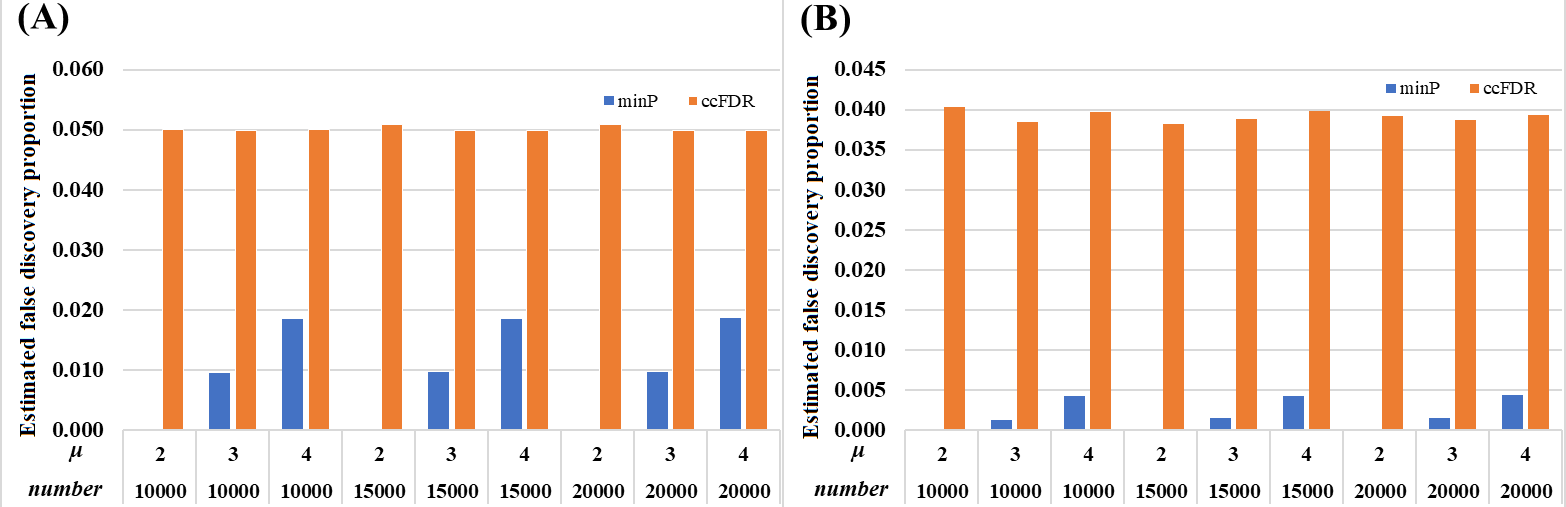

#### Figure S7. Estimated false discovery proportion for the minimum *P*-value method (minP) and cFDR with varying numbers of genetic loci (number=10000, 15000, and 20000). (A) Estimated false discovery proportion under the case with π_00_=0.40, π_10_=π_01_=0.20, and π_11_=0.20; (B) Estimated false discovery proportion control under the case with π_00_=0.80, π_10_=π_01_=0.05, and π_11_=0.10. These estimates were calculated by the average across 10^3^ replicates. In each case, *μ*_10_=*μ*_01_=*μ*_11_=*μ*=2. The significance level of FDR is set to 0.05.

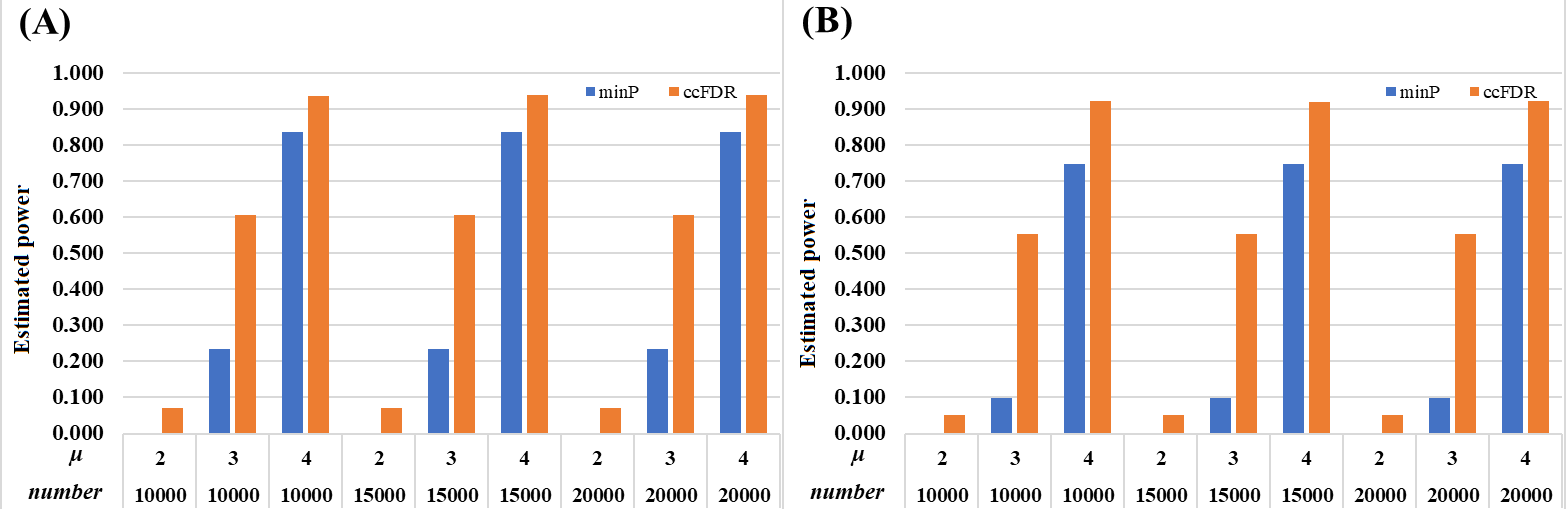

#### Figure S8. Estimated power for the minimum *P*-value method (minP) and cFDR with varying numbers of genetic loci (number=10000, 15000, and 20000). (A) Estimated false discovery proportion under the case with π_00_=0.40, π_10_=π_01_=0.20, and π_11_=0.20; (B) Estimated false discovery proportion control under the case with π_00_=0.80, π_10_=π_01_=0.05, and π_11_=0.10. These estimates were calculated by the average across 10^3^ replicates. In each case, *μ*_10_=*μ*_01_=*μ*_11_=*μ*=2. The significance level of FDR is set to 0.05.
